# Increased lipid peroxidation and lowered antioxidant defenses predict methamphetamine induced psychosis

**DOI:** 10.1101/2022.10.26.22281566

**Authors:** Hussein Kadhem Al-Hakeim, Mazin Fadhil Altufaili, Abbas F. Almulla, Shatha Rouf Moustafa, Michael Maes

**Affiliations:** Department of Chemistry, College of Science, University of Kufa, Kufa 54002, Iraq.,.; Department of Psychiatry, Faculty of Medicine, Chulalongkorn University, Bangkok, Thailand.; Medical Laboratory Technology Department, College of Medical Technology, The Islamic University, Najaf, Iraq.; Clinical Analysis Department, College of Pharmacy, Hawler Medical University, Erbil, Iraq..; Department of Psychiatry, Medical University of Plovdiv, Plovdiv, Bulgaria; Deakin University, IMPACT, the Institute for Mental and Physical Health and Clinical Translation, School of Medicine, Barwon Health, Geelong, Australia.

**Keywords:** psychosis, oxidative and nitrosative stress, antioxidants, neurotoxicity, schizophrenia

## Abstract

**Background:** A significant percentage of methamphetamine (MA) dependent patients develop psychosis. However, the associations between oxidative pathways and MA-induced psychosis (MIP) are not well delineated.

**Objective:** The aim of this study is to delineate whether acute MA intoxication in MA dependent patients is accompanied by increased nitro-oxidative stress and whether the latter is associated with MIP.

**Method:** We recruited 30 healthy males and 60 acutely intoxicated MA males with MA dependence and assessed severity of MA use and dependence and psychotic symptoms during intoxication and measured serum oxidative toxicity (OSTOX) biomarkers including oxidized high (oxHDL) and low (oxLDL)-density lipoprotein, myeloperoxidase (MPO), malondialdehyde (MDA), and nitric oxide (NO), and antioxidant defenses (ANTIOX) including HDL-cholesterol, zinc, glutathione peroxidase (GPx), total antioxidant capacity (TAC), and catalase-1.

**Results:** A large part (50%, n=30) of patients with MA dependence could be allocated to a cluster characterized by high psychosis ratings including delusions, suspiciousness, conceptual disorganization and difficulties abstract thinking and an increased OSTOX/ANTIOX ratio. Partial Least Squares analysis showed that 29.9% of the variance in MIP severity (a first factor extracted from psychosis, hostility, excitation, mannerism, and formal thought disorder scores) was explained by HDL, TAC and zinc (all inversely) and oxLDL (positively). MA dependence and dosing explained together 44.7% of the variance in the OSTOX/ANTIOX ratio.

**Conclusion:** MA dependence and intoxication are associated with increased oxidative stress and lowered antioxidant defenses, which both increase risk of MIP during acute intoxication. MA dependence is accompanied by increased atherogenicity due to lowered HDL and increased oxLDL and oxHDL.

## Introduction

Methamphetamine (MA), a potent psychostimulant derivative of amphetamine, is the second-most misused substance after cannabis, and is a worldwide health concern because of its ubiquity, high prevalence, and rising overdose-related death rates (Hogarth, Manning et al. 2021, Jones, Houry et al. 2022). In the United States, MA use increases among individuals who use other drugs, including heroin, from 22.5% to 46.7% in one year (Strickland, Stoops et al. 2021). People usually use MA to cope with their weariness, induce a state of pleasure, facilitate social interaction, enhance libido, boost productivity at home and work, and lose weight by decreasing appetite (Russell, Dryden et al. 2008, Shoptaw, Li et al. 2022). Nevertheless, several adverse consequences are associated with using MA, one of the most important being addiction (Beck, Larance et al. 2022, NIDA 2022). MA may trigger many neuropsychiatric symptoms and behaviors among users, including a wide spectrum of affective, cognitive, somatic, psychotic, and behavioral manifestations, including violent behavior, insomnia, and irritability (Meredith, Jaffe et al. 2005, McKetin, Lubman et al. 2014, Harro 2015, Wearne and Cornish 2018, May, Aupperle et al. 2020). Consequently, these symptoms may cause progressive social and occupational decline (Glasner-Edwards, Mooney et al. 2008, Glasner-Edwards, Mooney et al. 2010).

Many MA-dependent individuals may experience new-onset psychotic symptoms or a worsening of their psychotic symptoms (McKetin 2018). Auditory and tactile hallucinations, paranoid delusions, and ideas of reference are repeatedly reported as prominent MA-related psychotic symptoms (Zweben, Cohen et al. 2004, McKetin, McLaren et al. 2006). Previous studies indicate a wide prevalence range of MA-induced psychosis (MIP) ranging from 7% (McKetin, McLaren et al. 2006) to 76% (Salo, Fassbender et al. 2013). A meta-analysis revealed that the prevalence of MA-related psychotic disorders is 36.5%, with a lifetime prevalence of 42.7% (Lecomte, Dumais et al. 2018). Nevertheless, MIP is a challenging concept because the diagnostic criteria are not well-defined and because various etiologic and pathophysiological factors are associated with MIP. Patients with a genetic predisposition to psychosis and those who have already suffered from a psychotic disorder like schizophrenia are more likely to develop MA-associated psychotic symptoms (Batki and Harris 2004, McKetin, McLaren et al. 2006). Important risk factors for MIP are increased MA use, higher dependence, and frequent intake (Arunogiri, Foulds et al. 2018).

Schizophrenia and MA dependence and MIP share several clinical characteristics, and MIP is frequently considered to be a mechanistic model of schizophrenia (Ikeda, Okahisa et al. 2013, Gan, Song et al. 2018, Kalayasiri, Kraijak et al. 2019, Kalayasiri, Kraijak et al. 2019). Dopaminergic signaling in the mesocorticolimbic and nigrostriatal networks has been implicated in schizophrenia and MIP (Hogarth, Manning et al. 2021). The same disorders in LINE 1 partial methylation patterns are detected in MIP and paranoid schizophrenia and are more pronounced in the latter (Kalayasiri, Kraijak et al. 2019, Kalayasiri, Kraijak et al. 2019). Schizophrenia may result from neurotoxic processes (Maes, Sirivichayakul et al. 2020) and MA usage has neurotoxic effects on cortical interneurons (Hsieh, Stein et al. 2014). Activated immune-inflammatory and neuro-oxidative stress play a role not only in schizophrenia or schizophrenia phenotypes (Maes, Sirivichayakul et al. 2020, Ermakov, Dmitrieva et al. 2021, Cuenod, Steullet et al. 2022) but also in MA dependence (Limanaqi, Gambardella et al. 2018, Kalayasiri, Kraijak et al. 2019, Afzali, Fadaei et al. 2022). High levels of nitro-oxidative stress (NOS) are confirmed in schizophrenia as indicated by increased reactive oxygen (ROS) and nitrogen species (RNS), increased lipid peroxidation as indicated by increased levels of lipid hydroperoxides, and increased protein oxidation as indicated by increased advanced oxidation products (AOPP), and lowered total antioxidant defenses (Noto, Maes et al. 2015, Boll, Noto et al. 2017, Al-Hakeim, Almulla et al. 2020, Maes, Sirivichayakul et al. 2020, Roomruangwong, Noto et al. 2020). Oxidative stress biomarkers, including those indicating oxidative damage end-products, oxidant enzyme activities, and lowered antioxidant levels are also observed in MA dependent patients (Solberg, Refsum et al. 2019, Guidara, Messedi et al. 2020).

Therefore, we hypothesized that indicants of increased oxidative stress toxicity (OSTOX) and decreased antioxidant defenses (ANTIOX) may be detected in patients with MA dependence and MIP during MA intoxication. Nonetheless, no studies have reported associations between OSTOX/ANTIOX and MIP in MA-dependent individuals during MA intoxication. Hence, the aim of the present study is to examine whether MA dependence and MIP during intoxication are characterized by a) increased serum NOS/OSTOX biomarkers, including MDA, myeloperoxidase (MPO), nitric oxide (NO), oxidized high-density lipoprotein (oxHDL) and low-density lipoprotein (oxLDL) levels; and b) lowered ANTIOX biomarkers, including catalase-1, glutathione peroxidase (GPx), total antioxidant capacity (TAC), HDL cholesterol, and zinc. The data is analyzed using a precision nomothetic approach (Maes 2022).

## Material and Methods

### Participants

In the present study, we recruited sixty MA-intoxicated male patients with MA substance use disorder (SUD) at the Psychiatry Unit, Al-Hussein Medical City, Kerbala Governorate, Iraq, from April 2022 to August 2022. The patients were diagnosed according to the Diagnostic and Statistical Manual of Mental Disorders (5th edition) (DSM-5) as moderate to severe SUD (American Psychiatric Association 2013). Because of the religious state of Karbala city, we could not recruit female MA SUD patients and included male SUD patients only. All SUD patients started MA intake before at least three months prior to the study. A urine examination showed a positive MA test. Thirty apparently healthy controls, namely family and friends of staff or friends of patients served as controls. None of the controls had ever taken any psychoactive drugs (except tobacco use disorder) and none showed a current or lifetime DSM-5 axis I diagnosis of SUD, psychosis or schizophrenia, bipolar disorder or other affective disorders, or a family history in first-degree relatives of schizophrenia or psychosis. Patients and controls were excluded if they had ever taken immunosuppressive treatments or glucocorticoids or had been diagnosed with a neurodegenerative or neuroinflammatory illness such as Alzheimer’s disease, Parkinson’s disease, multiple sclerosis, or stroke. Additionally, individuals with (auto)immune diseases such as inflammatory bowel disease, rheumatoid arthritis, COPD, psoriasis, or diabetes mellitus were excluded.

The study followed Iraqi and international privacy and ethics laws. Before participating in this study, all participants, and their guardians (legal representatives of the patients are mother, father, brother, spouse, or son) gave written informed consent. The study was approved by the ethics committee (IRB) of the College of Science, University of Kufa, Iraq (89/2022), Karbala Health Directorate-Training and Human Development Center (Document No.18378/ 2021), which follows the Declaration of Helsinki’s International Guideline for Human Research Protection.

### Clinical assessments

To collect patient and control data, a senior psychiatrist with expertise in addiction conducted a semi-structured interview and scored rating scales to assess severity of MA dependence, use and psychosis. The Severity of the Dependence Scale (SDS) was used to estimate the severity of MA dependence, namely 5 items: a) did you ever think your use of MA was out of control; b) did the prospect of missing MA make you very anxious or worried; c) did you worry about your use of MA; d) did you wish you could stop; and e) how difficult would you find it to stop or go without MA (Gossop, Darke et al. 1995). We also registered age at onset, duration of MA dependence, daily dosage (grams), route of administration (ordinal variable with no=0, orally ingested=1, smoked or snorted=2, and injected=3), number of previous psychotic episodes and days hospitalized after admission for acute intoxication. We also registered lifetime cannabis and alcohol use as well as cannabis and alcohol dependence. The patients did not use any other drugs of dependence, including opioids, cocaine, or heroin. The same day, we assessed the Brief Psychiatric Rating Scale (BPRS) (Overall and Gorham 1962) and the Positive and Negative Syndrome Scale (PANSS) (Kay, Fiszbein et al. 1987) rating scales. In analogy with our studies in schizophrenia (Al-Dujaili, Mousa et al. 2021, Almulla, Al-Hakeim et al. 2021, Maes, Vojdani et al. 2021), we computed different z unit-weighted composite scores based on the items of the BPRS and PANNS to reflect the severity of MIP, namely: (a) psychosis as sum of the z transformations of hallucinations (BPRS) + suspiciousness (BPRS) + delusions (PANSP1) + hallucinatory behavior (PANSS P3) + suspiciousness (PANNS P6); (b) hostility was computed as the sum of z transformations of hostility (BPRS) + uncooperativeness (BPRS) + hostility (PANSS P7) + uncooperativeness (PANNS G8) + poor impulse control (PANSS G14); (c) excitement: sum of z transformations of grandiosity (BPRS) + excitement (BPRS) + excitement (PANNS P4) + grandiosity (PANNS P5), (d) mannerism and posturing (BPRS) + mannerism and posturing (PANNS G5); (e) formal thought disorders (FTD): sum of z transformations of conceptual disorganization (BPRS) + unusual thoughts (BPRS) + conceptual disorganization (PANNS P2) + difficulties in abstract thinking (PANNS N5) + stereotyped thinking (PANNS N7). In addition, we computed, post-hoc, a new index based on the most prominent psychosis-associated MA intoxication (MAI) symptoms in our MA dependent patients, namely delusions (PANSS P1) + conceptual disorganization (PANSS P2) + suspiciousness (PANNS P6) + difficulty in abstract thinking (PANNS N5). Tobacco use disorder (TUD) was diagnosed following DSM-5 criteria. The following formula was used to compute the body mass index (BMI): body weight (kg) / length (m^2^).

### Biomarkers assays

Fasting venous blood was obtained from all participants in the early morning hours after awakening and before having breakfast. After 15 minutes at room temperature, blood was allowed to coagulate for 10 minutes before being centrifuged at 3,000 rpm for 10 minutes. Separated serum was then transferred to Eppendorf tubes and stored at −80 °C until analysis. A urine MA test was done immediately after admitting the acutely intoxicated patient using the urine Multi-Drug 12 Drugs Rapid Test Panel kit supplied by Citest Diagnostics Inc. (Vancouver, Canada). Serum zinc was measured spectrophotometrically using a ready-for-use kit supplied by Agappe Diagnostics^®^ (Cham, Switzerland). HDL was measured using a kit supplied by Spinreact® (Gerona, Spain) based on a direct method. Serum levels of catalase, Gpx, MPO, MDA, oxHDL, oxLDL, TAC, and NO were measured using commercial ELISA kits supplied by Nanjing Pars Biochem Co. Ltd. (Nanjing, China). All kits were based on a sandwich technique. The procedures were followed according to the manufacturer’s instructions without any modifications. The intra-assay coefficients of variation (CV) for all the assays were <10.0% (precision within-assay). Consequently, we computed 3 composite scores: a) oxidative stress toxicity (OSTOX) as the sum of the z transformation of MPO (zMPO) + zMDA + zoxHDL + zoxLDL; b) antioxidant defenses (ANTIOX): zcatalase + zGpx + zTAC + zZn + zHDL; and the OSTOX/ANTIOX ratio as zOSTOX – zANTIOX.

### Statistical analysis

Analysis of variance (ANOVA) was employed to examine differences between groups in continuous variables and analysis of contingency tables (χ2-test) to investigate the association between nominal variables. Pearson’s product-moment correlation coefficient was employed to examine the correlation between two scale variables. We utilized a multivariate general linear model (GLM) to delineate the associations between study group (healthy control and patient groups) and the psychiatric rating scale scores composites and biomarkers, while controlling for confounding variables, namely age, sex, smoking, and education. We calculated the estimated marginal mean values (SE) and employed protected (namely: the omnibus test is significant) LSD tests to carry out pairwise comparisons among the group means. We additionally applied false discovery rate (FDR) p-correction to the multiple comparisons (Benjamini and Hochberg 1995). In addition, multiple regression analysis has been utilized to examine whether the biomarkers can significantly predict the various symptom domains. We also used a stepwise automated approach with a p-value of 0.05 for entry and 0.06 for removal from the model. Standardized beta coefficients with t statistics and exact p-value were computed for each of the predictors’ variables and we also compute the model statistics (F, df and p values) and total variance explained (R^2^) as effect size. Furthermore, we used the variance inflation factor (VIF) and tolerance to examine collinearity and multicollinearity issues. We tested for heteroskedasticity using the White and modified Breusch-Pagan homoscedasticity tests and, if necessary, utilized univariate GLM analysis to estimate parameters with substantial error margins. Two-tailed tests were used to evaluate the significance, set at p=0.05 in SPSS version 28 (windows) to perform all statistics.

Partial Least Squares (PLS) analysis was conducted to delineate the causative associations among MA dependence, alterations in ONS biomarkers, and the symptom domain scores induced by MA. Toward this end, we examined if one validated latent vector could be extracted from psychosis, hostility, excitement, and formal thought disorders and, if so, used this factor as output variable. The biomarker input variables were entered as single indicators and the common input variable was a latent vector reflecting MA-dependence and use. We perform complete PLS analysis when the following criteria are met: a) all loadings on the latent vectors should be > 0.6 at p < 0.001, b) adequate construct and convergence validity as indicated by rho A > 0.8, Cronbach’s alpha > 0.7, composite reliability > 0.7, and average variance extracted (AVE) > 0.5, c) construct’s cross-validated redundancy should be sufficient as indicated by blindfolding analysis, d) the results of the Confirmatory Tetrad Analysis (CTA) should display that the latent vectors constructed are correctly described as a reflective model, e) PLSpredict analysis should prove that prediction performance of the model is efficient, and f) adequate model fit as indicated by standardized root squared residual (SRMR) values < 0.08. Once the quality of the model has been confirmed based on the criteria mentioned above, we carried out a complete PLS-SEM pathway analysis using 5,000 bootstraps to compute the path coefficients (with p-values) along with specific and total indirect (mediated) effects and total effects. A priori power analysis shows that the estimated sample size for a PLS analysis (which is the primary analysis) performed using a power = 0.8, alpha =0.05, effect size = 0.17 and using maximal 5 predictors should be 82 participants. Accordingly, we included 90 subjects in the present study.

## Results

### Cluster and factor analysis

We performed a two-step cluster analysis to divide the patients into two groups according to the MAI symptoms, OSTOX, ANTIOX and OSTOX/ANTIOX, while we entered MA dependent patients versus controls as categorical variable. Three clusters were formed with a silhouette measure of cohesion and separation of 0.62. These included healthy controls (n=30) and individuals with lower psychotic scores and lower NOS (MA-PSO, n=30) versus those with high psychotic scores and high NOS (MA+PSO, n=30). We were able to extract validated PCs from SDS1 (loading=0.913), SDS2 (0.951), SDS4 (and 0.691), and SDS5 (0.878) (KMO=0.832, Bartlett’s test chi-square=287.09, df=6, p<0.001, AVE=0.747, labeled PC_SDS). We were also able to extract a validated PC from PC_SDS (0.959), dosage (0.854), MA use last month (0.961) and route of administration entered as an ordinal variable (0.672) (KMO=0.743, Bartlett’s test chi-square=367.85, df=6, p<0.001, AVE=0.756, labelled: PC_MA).

### Sociodemographic data and MA features in the study groups

The sociodemographic characteristics of controls and both MA subgroups are presented in **Table 1**. The results show that MA+PSO patients are older and show a higher unemployment rate and PC_SDS and PC-MA scores than MA-PSO patients, whilst there are no significant differences in BMI, education, and marital state. Other differences between both MA groups are a higher rate of injections of abused MA, duration of MA dependence, and dosing of MA in the MA+PSO group than in the MA-PSO group, whilst there were no differences in number of MIP episodes, duration of index MIP episode, and days admitted to hospital due to MA intoxication. MA abusing patients show more TUD than controls but no differences in alcohol dependence and current intake or lifetime cannabis use. None of the patients or controls showed any abuse of other illicit drugs including cocaine, heroin, or opioids.

**Table 1.** Socio-demographic and methamphetamine abuse (MA) data in healthy control participants (HCP) and MA patients classified as those with (MA+PSO) and without (MA-PSO) increased psychotic symptoms and oxidative stress biomarkers

| Variables | HCP (n=30)<sup>A</sup> | MA-PSO (n=30)<sup>B</sup> | MA+PSO (n=30)<sup>C</sup> | F/X<sup>2</sup> | df | p |
| --- | --- | --- | --- | --- | --- | --- |
| Age (years) | 27.3 (5.4) | 24.4 (6.6) <sup>C</sup> | 28.6 (5.4) <sup>B</sup> | 3.99 | 2/87 | 0.022 |
| BMI (Kg/m2) | 25.33 (2.80) | 25.24 (4.05) | 24.94 (3.12) | 0.11 | 2/87 | 0.894 |
| Education (years) | 10.9 (3.7) | 7.8 (7.0) | 10.2 (6.8) | 2.16 | 2/87 | 0.121 |
| Marital state (Single/Married) | 8/22 | 13/17 | 8/22 | 2.54 | 2 | 0.280 |
| Employment (No/Yes) | 8/22 <sup>C</sup> | 10/20 <sup>C</sup> | 25/5 <sup>A,B</sup> | 23.07 | 2 | <0.0001 |
| Current MA use (No/Yes) | 0/30 | 30/0 | 30/0 | 90.0 |  |  |
| PC_SDS (z score) | -1.370 (0.0) <sup>B,C</sup> | 0.611 (0.297) <sup>A,C</sup> | 0.758 (0.237) <sup>A,B</sup> | KWT |  | <0.001 |
| PC_MA dependence severity (z score) | -1.382 (0.0) <sup>B,C</sup> | 0.570 (0.166) <sup>A,C</sup> | 0.812 (0.219) <sup>A,B</sup> | KWT |  | <0.0001 |
| MA Administration route (O/S/I) | - | 4/16/7 | 13/5/12 | FFHET |  | 0.001 |
| Age at onset (years) | - | 23.4 (5.9) <sup>C</sup> | 25.8 (4.90) <sup>B</sup> | 3.02 | 1/58 | 0.088 |
| Duration of MA dependence (months) | - | 14.8 (14.1) <sup>C</sup> | 34.9 (17.2) <sup>B</sup> | 24.54 | 1/57 | <0.001 |
| MA dosing (gm) | - | 1.13 (0.47) <sup>C</sup> | 2.30 (0.79) <sup>B</sup> | 47.81 | 1/58 | <0.001 |
| Number of prior MIP episodes | - | 2.0 (2.4) | 2.2 (1.4) | 0.07 | 1/58 | 0.798 |
| Duration of the index MIP (days) | - | 2.4 (2.3) <sup>A</sup> | 2.8 (2.0) <sup>A</sup> | 0.43 | 1/58 | 0.513 |
| Days hospitalized due to MA intoxication | - | 1.3 (1.7) <sup>A</sup> | 1.8 (1.8) <sup>A</sup> | 1.19 | 1/58 | 0.280 |
| TUD (No/Yes) | 15/15 | 7/23 | 1/29 | 17.29 | 2 | <0.001 |
| Alcohol dependence (No/Yes) | 30/0 | 29/1 | 28/2 | FFHET |  | 0.770 |
| Current drinker/past month (No/Yes) | 30/0 | 29/1 | 28/2 | FFHET |  | 0.770 |
| Lifetime Cannabis use (No/Yes) | 30/0 | 29/1 | 28/2 | FFHET |  | 0.770 |
| Any other substance, dependence | 0 | 0 | 0 | - | - | - |

Results are shown as mean (SD) or as ratios: F: results of analysis of variance; $X^2$ : analysis of contingency tables.
BMI: Body Mass Index, O/S/: orally, smoking, injection, Kg: Kilogram, gm: Gram, TUD: Tobacco use disorder
<sup>A,B,C</sup>: Results of pairwise comparisons among group means.

### Psychotic symptoms scores among study groups

**Table 2** shows the measurements of MIP associated symptoms, namely MAI symptoms, psychosis, hostility, excitement, mannerism and FTD, in the three study groups. There were significant differences in all MIP-associated domains between MA-PSO and MA+PSO, except in mannerism. These results remained significant after FDR correction.

**Table 2.**
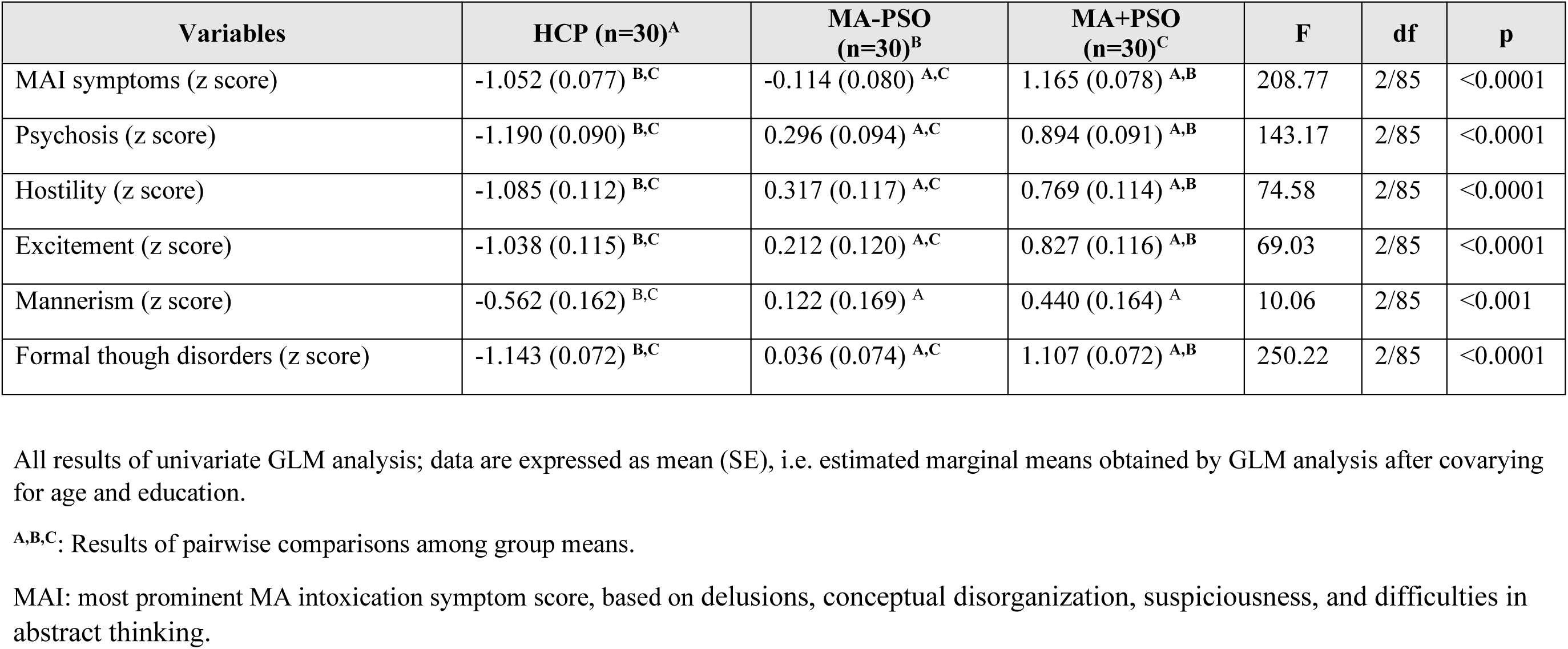
Clinical rating scales scores in healthy control participants (HCP) and patients with methamphetamine abuse (MA) classified into those with (MA+PSO) and without (MA-PSO) increased psychotic symptoms and oxidative stress (PSO)

| Variables | HCP (n=30)<sup>A</sup> | MA-PSO (n=30)<sup>B</sup> | MA+PSO (n=30)<sup>C</sup> | F | df | p |
| --- | --- | --- | --- | --- | --- | --- |
| MAI symptoms (z score) | -1.052 (0.077) <sup>B,C</sup> | -0.114 (0.080) <sup>A,C</sup> | 1.165 (0.078) <sup>A,B</sup> | 208.77 | 2/85 | <0.0001 |
| Psychosis (z score) | -1.190 (0.090) <sup>B,C</sup> | 0.296 (0.094) <sup>A,C</sup> | 0.894 (0.091) <sup>A,B</sup> | 143.17 | 2/85 | <0.0001 |
| Hostility (z score) | -1.085 (0.112) <sup>B,C</sup> | 0.317 (0.117) <sup>A,C</sup> | 0.769 (0.114) <sup>A,B</sup> | 74.58 | 2/85 | <0.0001 |
| Excitement (z score) | -1.038 (0.115) <sup>B,C</sup> | 0.212 (0.120) <sup>A,C</sup> | 0.827 (0.116) <sup>A,B</sup> | 69.03 | 2/85 | <0.0001 |
| Mannerism (z score) | -0.562 (0.162) <sup>B,C</sup> | 0.122 (0.169) <sup>A</sup> | 0.440 (0.164) <sup>A</sup> | 10.06 | 2/85 | <0.001 |
| Formal thought disorders (z score) | -1.143 (0.072) <sup>B,C</sup> | 0.036 (0.074) <sup>A,C</sup> | 1.107 (0.072) <sup>A,B</sup> | 250.22 | 2/85 | <0.0001 |

All results of univariate GLM analysis; data are expressed as mean (SE), i.e. estimated marginal means obtained by GLM analysis after covarying for age and education.
<sup>A,B,C</sup>: Results of pairwise comparisons among group means.
MAI: most prominent MA intoxication symptom score, based on delusions, conceptual disorganization, suspiciousness, and difficulties in abstract thinking.

### Serum biomarkers levels among the study groups

The measured biomarkers are presented in **Table 3**. The results show that Gpx, NO, and zinc are significantly decreased in MA+PSO as compared with controls. MDA, oxLDL and OSTOX (all three increased) and HDL (decreased) were significantly different between MA patients and controls. TAC and ANTIOX and the OSTOX/ANTIOX ratio were significantly different between the three study groups, with TAC and ANTIOX decreasing and OSTOX/ANTIOX increasing from controls → MA-PSO → MA+PSO. oxHDL was significantly higher in the MA+PSO group than in controls.

**Table 3.**
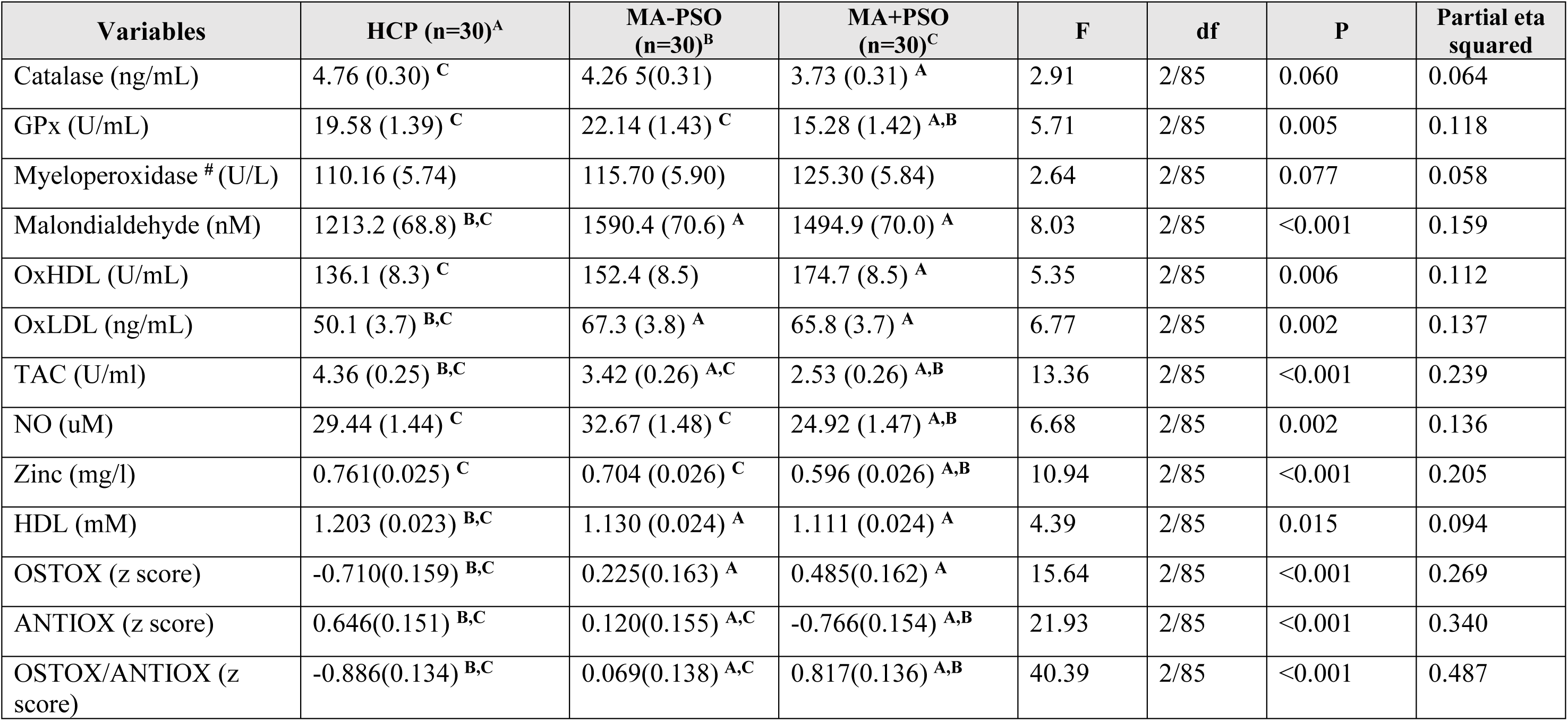
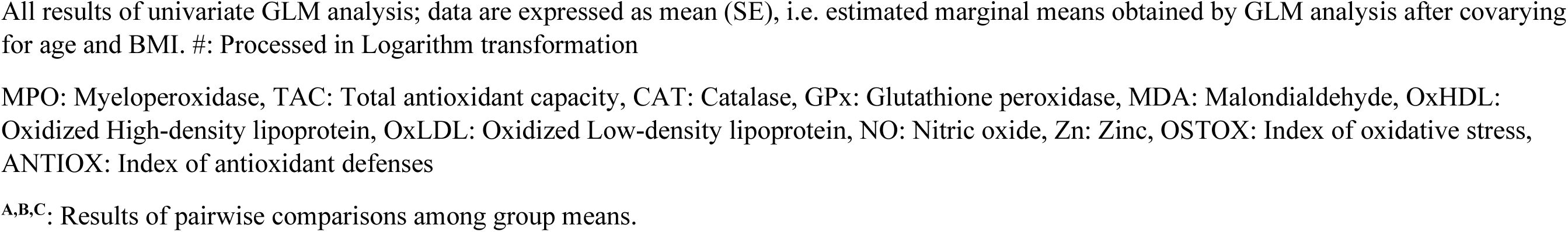
Oxidative stress biomarkers in heathy control participants (HCP) and patients with methamphetamine abuse (MA) classified into those with (MA+PSO) and without (MA-PSO) increased psychotic symptoms and oxidative stress (PSO)

| Variables | HCP (n=30)<sup>A</sup> | MA-PSO (n=30)<sup>B</sup> | MA+PSO (n=30)<sup>C</sup> | F | df | P | Partial eta squared |
| --- | --- | --- | --- | --- | --- | --- | --- |
| Catalase (ng/mL) | 4.76 (0.30) <sup>C</sup> | 4.26 5(0.31) | 3.73 (0.31) <sup>A</sup> | 2.91 | 2/85 | 0.060 | 0.064 |
| GPx (U/mL) | 19.58 (1.39) <sup>C</sup> | 22.14 (1.43) <sup>C</sup> | 15.28 (1.42) <sup>A,B</sup> | 5.71 | 2/85 | 0.005 | 0.118 |
| Myeloperoxidase <sup>#</sup> (U/L) | 110.16 (5.74) | 115.70 (5.90) | 125.30 (5.84) | 2.64 | 2/85 | 0.077 | 0.058 |
| Malondialdehyde (nM) | 1213.2 (68.8) <sup>B,C</sup> | 1590.4 (70.6) <sup>A</sup> | 1494.9 (70.0) <sup>A</sup> | 8.03 | 2/85 | <0.001 | 0.159 |
| OxHDL (U/mL) | 136.1 (8.3) <sup>C</sup> | 152.4 (8.5) | 174.7 (8.5) <sup>A</sup> | 5.35 | 2/85 | 0.006 | 0.112 |
| OxLDL (ng/mL) | 50.1 (3.7) <sup>B,C</sup> | 67.3 (3.8) <sup>A</sup> | 65.8 (3.7) <sup>A</sup> | 6.77 | 2/85 | 0.002 | 0.137 |
| TAC (U/ml) | 4.36 (0.25) <sup>B,C</sup> | 3.42 (0.26) <sup>A,C</sup> | 2.53 (0.26) <sup>A,B</sup> | 13.36 | 2/85 | <0.001 | 0.239 |
| NO (uM) | 29.44 (1.44) <sup>C</sup> | 32.67 (1.48) <sup>C</sup> | 24.92 (1.47) <sup>A,B</sup> | 6.68 | 2/85 | 0.002 | 0.136 |
| Zinc (mg/l) | 0.761(0.025) <sup>C</sup> | 0.704 (0.026) <sup>C</sup> | 0.596 (0.026) <sup>A,B</sup> | 10.94 | 2/85 | <0.001 | 0.205 |
| HDL (mM) | 1.203 (0.023) <sup>B,C</sup> | 1.130 (0.024) <sup>A</sup> | 1.111 (0.024) <sup>A</sup> | 4.39 | 2/85 | 0.015 | 0.094 |
| OSTOX (z score) | -0.710(0.159) <sup>B,C</sup> | 0.225(0.163) <sup>A</sup> | 0.485(0.162) <sup>A</sup> | 15.64 | 2/85 | <0.001 | 0.269 |
| ANTIOX (z score) | 0.646(0.151) <sup>B,C</sup> | 0.120(0.155) <sup>A,C</sup> | -0.766(0.154) <sup>A,B</sup> | 21.93 | 2/85 | <0.001 | 0.340 |
| OSTOX/ANTIOX (z score) | -0.886(0.134) <sup>B,C</sup> | 0.069(0.138) <sup>A,C</sup> | 0.817(0.136) <sup>A,B</sup> | 40.39 | 2/85 | <0.001 | 0.487 |

All results of univariate GLM analysis; data are expressed as mean (SE), i.e. estimated marginal means obtained by GLM analysis after covarying for age and BMI. #: Processed in Logarithm transformation
MPO: Myeloperoxidase, TAC: Total antioxidant capacity, CAT: Catalase, GPx: Glutathione peroxidase, MDA: Malondialdehyde, OxHDL: Oxidized High-density lipoprotein, OxLDL: Oxidized Low-density lipoprotein, NO: Nitric oxide, Zn: Zinc, OSTOX: Index of oxidative stress, ANTIOX: Index of antioxidant defenses
<sup>A,B,C</sup>: Results of pairwise comparisons among group means.

We also computed the differences in biomarkers between MA dependent patients and controls. We found that MDA, oxLDL OSTOX and OSTOX/ANTIOX ratio (all p<0.001) and oxHDL (p=0.012) were significantly higher in MA dependence than in controls, while catalase (p=0.033), HDL (p=0.005), TAC, zinc and ANTIOX (all p<0.001) were significantly lower in MA dependence than in controls. FDR p correction did not change any of these results.

### Intercorrelation between PC_SDS, PC_MA, biomarkers, and psychotic symptoms

We performed correlation analyses to delineate the associations between PC_SDS, PC_MA, MA-induced psychotic symptoms, and biomarkers (**Table 4).** Our results indicate that PC_SDS and PC_MA are significantly associated with all MA-induced symptom domains in both groups combined. Moreover, also in MA patients there were significant correlations between PC_SDS and PC_MA and MAI symptoms, psychosis, excitement and FTD. OSTOX (positively), ANTIOX (inversely) and OSTOX/ANTIOX (positively) were significantly associated with all MIP symptom domains in both controls and patients combined, except ANTIOX which was not significantly associated with mannerism. In MA patients, no significant correlations were detected between the OSTOX and ANTIOX scores and the symptom domains.

**Table 4.** Intercorrelations between severity of methamphetamine (MA) dependence (PC_SDS) and severity of MA dependence (PC_MA), methamphetamine abuse (MA), oxidative stress parameters, and symptoms domains.

| Variables | All subjects combined (n=90) |  |  |  |  | MA dependence (n=60) |  |
| --- | --- | --- | --- | --- | --- | --- | --- |
|  | PC_SDS | PC_MA | OSTOX | ANTIOX | OSTOX/ANTIOX | PC_SDS | PC_MA |
| MAI symptoms | 0.783** | 0.799** | 0.434** | -0.415** | 0.555** | 0.330* | 0.483** |
| Psychosis | 0.873** | 0.871** | 0.491** | -0.402** | 0.583** | 0.498** | 0.524** |
| Hostility | 0.790** | 0.793** | 0.360** | -0.285* | 0.422** | 0.189 | 0.201 |
| Excitement | 0.782** | 0.779** | 0.336** | -0.269* | 0.396** | 0.409** | 0.428** |
| Mannerism | 0.453** | 0.477** | 0.220* | -0.109 | 0.215* | 0.110 | 0.075 |
| Formal Thought Disorders | 0.839** | 0.850** | 0.451** | -0.426** | 0.573** | 0.382** | 0.500** |
| OSTOX | 0.520** | 0.520** | - |  |  | 0.164 | 0.168 |
| ANTIOX | -0.443** | -0.482** |  |  |  | 0.049 | -0.169 |
| OSTOX/ANTIOX ratio | 0.629** | 0.654** |  |  |  | 0.080 | 0.243 |

\*p<0.05; \*\*p<0.001
MAI: most prominent MA-intoxication symptoms, based on delusions, conceptual disorganization, suspiciousness, and difficulties in abstract thinking. OSTOX: index of oxidative toxicity; ANTIOX: index of antioxidant defenses

### Prediction of MIP symptoms and the OSTOX/ANTIOX ratio

In **Table 5,** regression #1, we selected MAI symptoms as a dependent variable and performed multiple regression analysis showing that 35.2% of the variance could be explained by TAC, HDL and zinc (inversely) and oxHDL and oxLDL (positively associated). Forced entry of age, BMI, education, TUD, and alcohol dependence showed that TUD (t=-2.14, p=0.035) was the only significant predictor and that the effect of the biomarkers remained significant. **Table 5,** regression #2 shows that 30.8% of the variance in the MAI score was explained by the OSTOX/ANTIOX ratio. **Figure 1** shows the partial regression of the MAI symptom score on the OSTOX/ANTIOX ratio (adjusted for age, TUD, education, BMI, and alcohol use). Adding TUD, showed that both the OSTOX/ANTIOX ratio and TUD were significant predictors and together explained 35.9% of the variance (F=24.34, df=2/89, p<0.001), although the impact of OSTOX/ANTIOX (β=0.479, t=5.59, p<0.001) was much higher than that of TUD (β=0.238, t=2.63, p=0.010). Nevertheless, univariate GLM analysis shows that TUD (F=0.017, df=1/84, p=0.896) has no significant effect on the MAI score above and beyond that of the diagnostic classification (F=66.53, df=2/84, p<0.001).

**Table 5.**
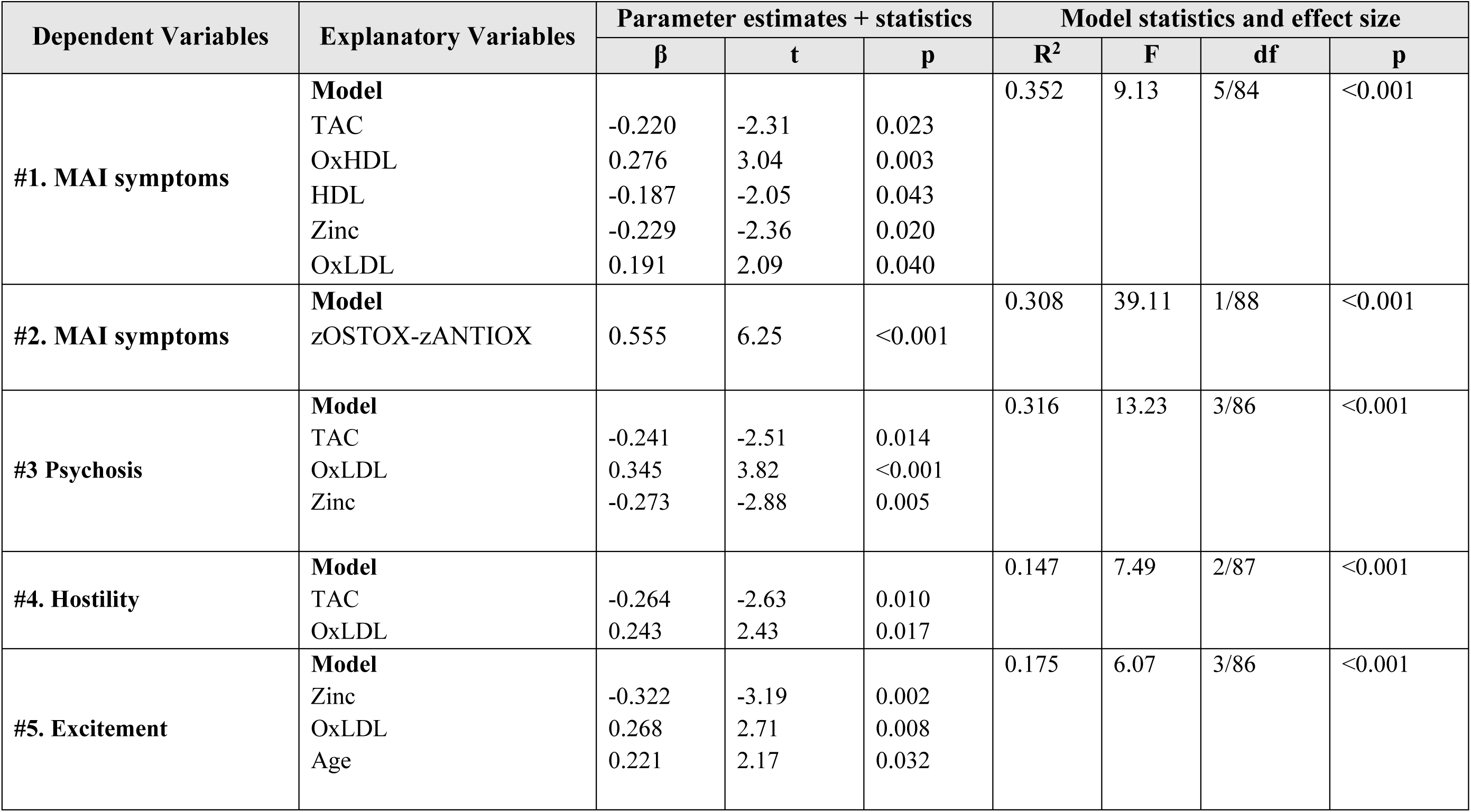
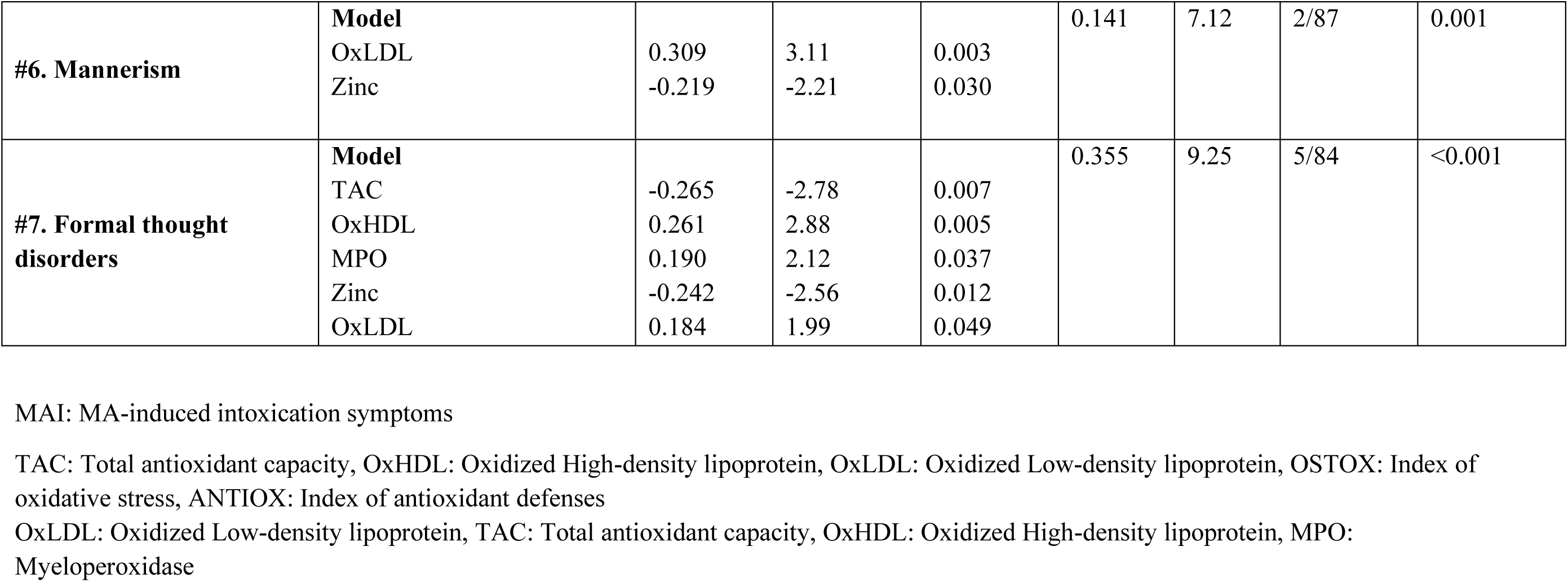
Results of multiple regression analyses with the methamphetamine (MA) intoxication symptoms as dependent variables and oxidative stress biomarkers as explanatory variables

**Figure 1.**
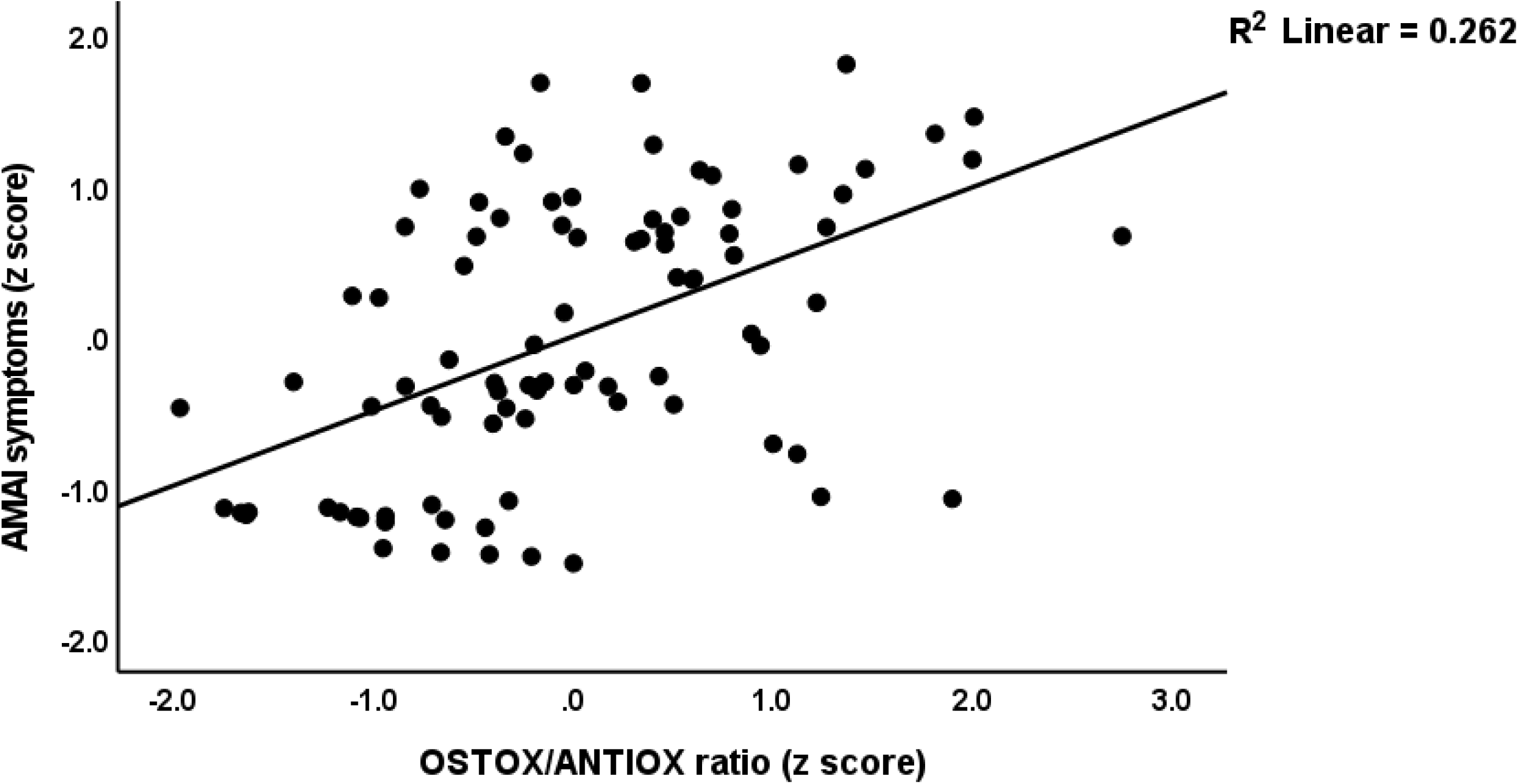
Partial regression plot of the methamphetamine intoxication (MAI) symptom score on the oxidative stress toxicity / antioxidant (OSTOX/ANTIOX) ratio

The second part of Table 5 shows the results of multiple regression analyses with MA-induced psychosis, hostility, excitement, mannerism and formal thought disorders as dependent variables and OSTOX and ANTIOX biomarkers as explanatory variables, while allowing for the effects of confounders. **Table 5**, regression #3, indicates that 31.6% of the variance in psychosis was explained by the regression on TAC and zinc (inversely) and oxLDL (positively). Regression #4 shows that TAC (inversely) and oxLDL (positively) could explain 14.7% of the variance in hostility. Regression #5 reveals that 17.5% of the variance in the excitement could be explained by zinc (inversely) and oxLDL and age (both positively). **Figure 2** shows the partial regression of the excitement score on serum zinc. Regression #6 indicates that 14.1% of the variance in mannerism was explained by the cumulative effects of oxLDL (positively) and zinc (inversely). A larger part of the variance in FTD (35.5%) was explained by the regression on TAC and zinc (both inversely), oxHDL and MPO (both positively). **Figure 3** shows the partial regression of the FTD score on oxHDL.

**Figure 2.**
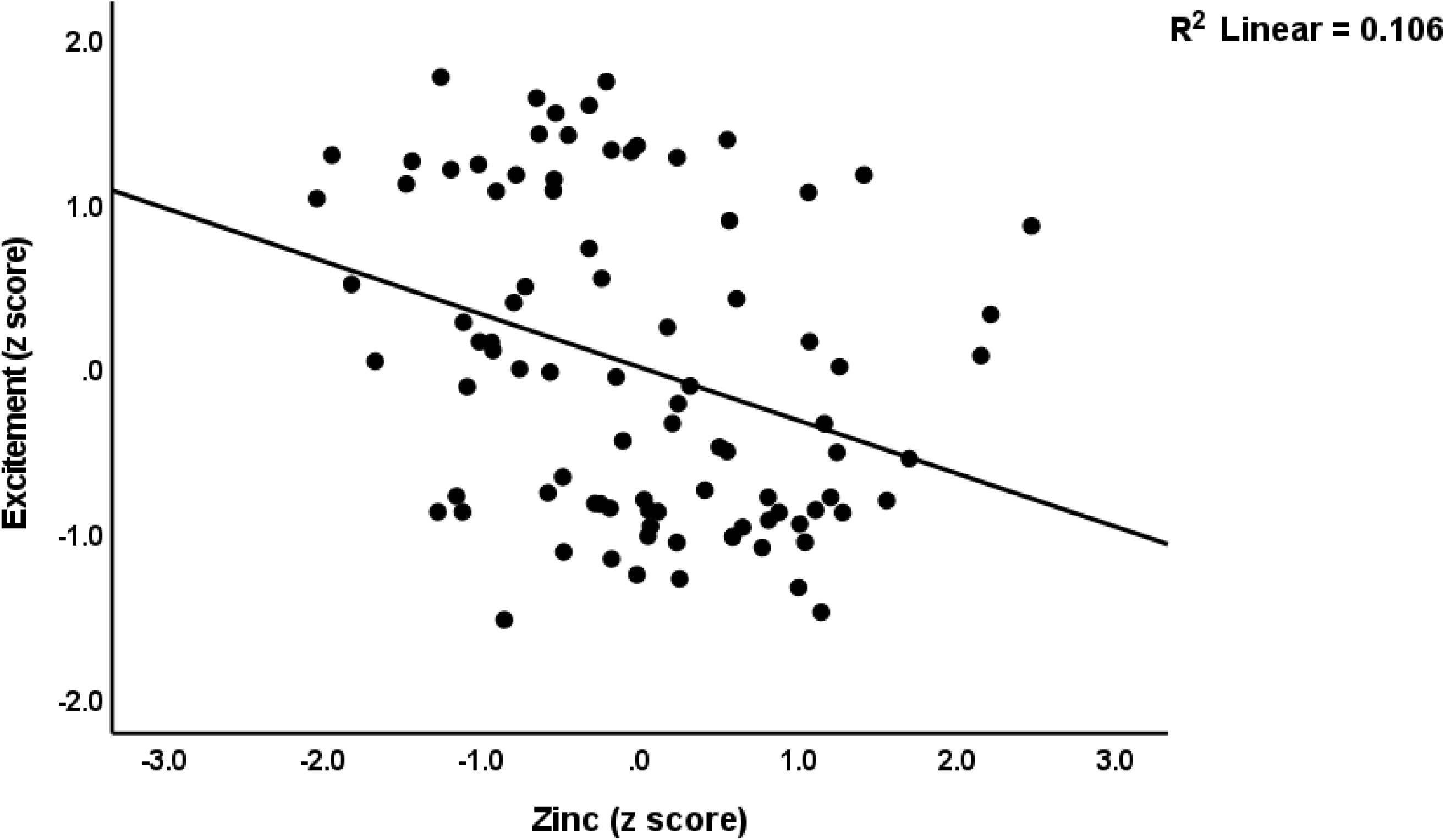
Partial regression plot of excitement on serum zinc levels

**Figure 3.**
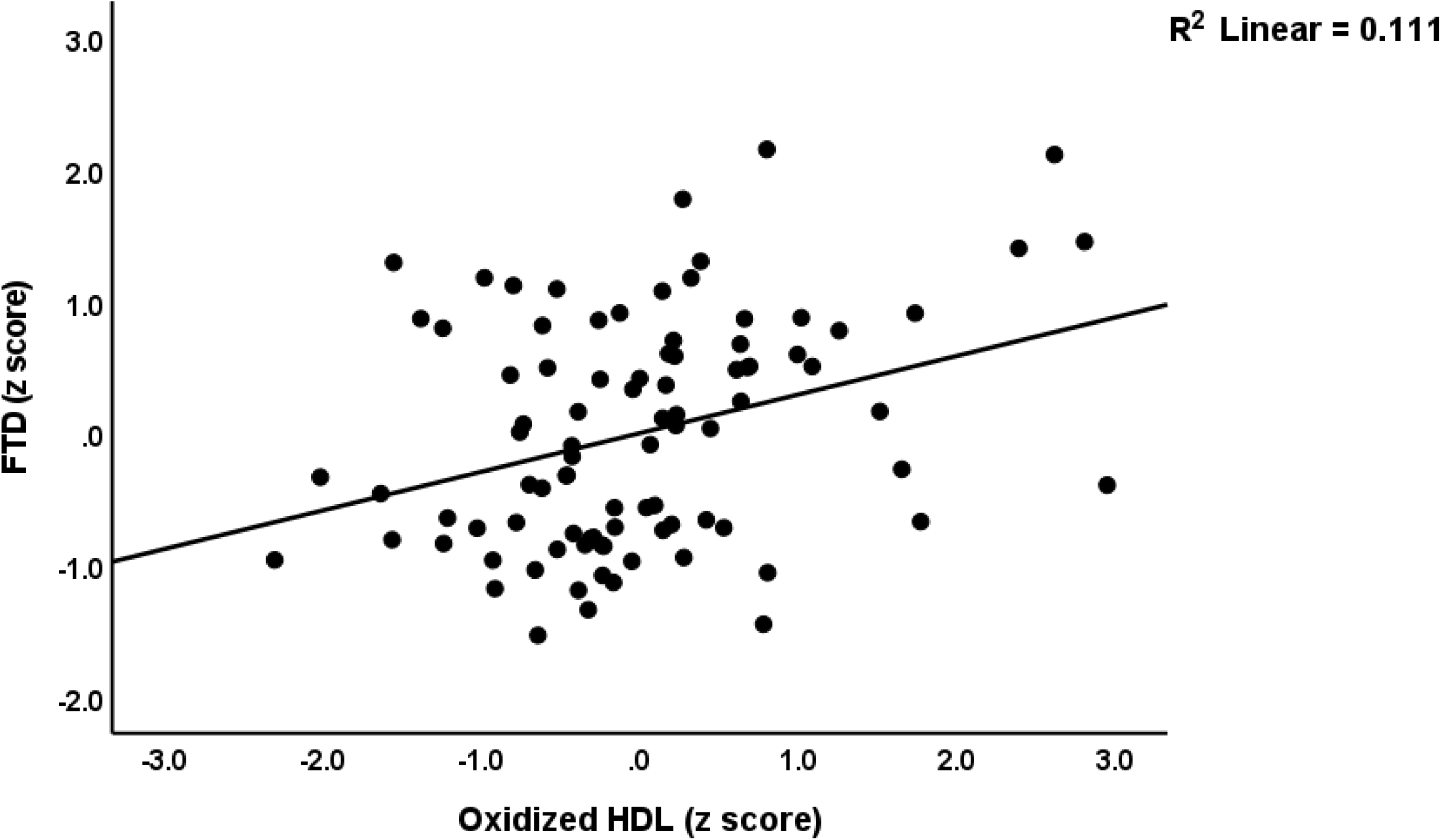
Partial regression plot of formal thought disorders (FTD) on serum oxidized high density lipoprotein (HDL) levels

Multiple regression analysis (**Table 6**, regression #1) showed that PC_SDS and MA dosing were the most significant predictors of the first PC extracted from the 5 symptom domains and explained 81.8% of its variance. **Figure 4** shows the partial regression of MA symptoms on PC_SDS. Multiple regression analysis (**Table 6**, regression #2) showed that PC_SDS and MA dosing were the most significant predictors of the OSTOX/ANTIOX ratio and explained 44.7% of its variance. **Figure 5** shows the partial regression of the OSTOX/ANTIOX ratio on MA dosing. In MA patients, we found that 7.1% of the variance in the OSTOX/ANTIOX ratio was explained by MA dosing (β=0.267, t=0.211, p=0.039). We found that 27.1% of the variance in OSTOX was explained by PC_SDS (F=32.64, df=1/88, p<0.001), while 21.6% in ANTIOX was explained by MA dosing (F=24.22, df=1/88, p<0.001).

**Figure 4.**
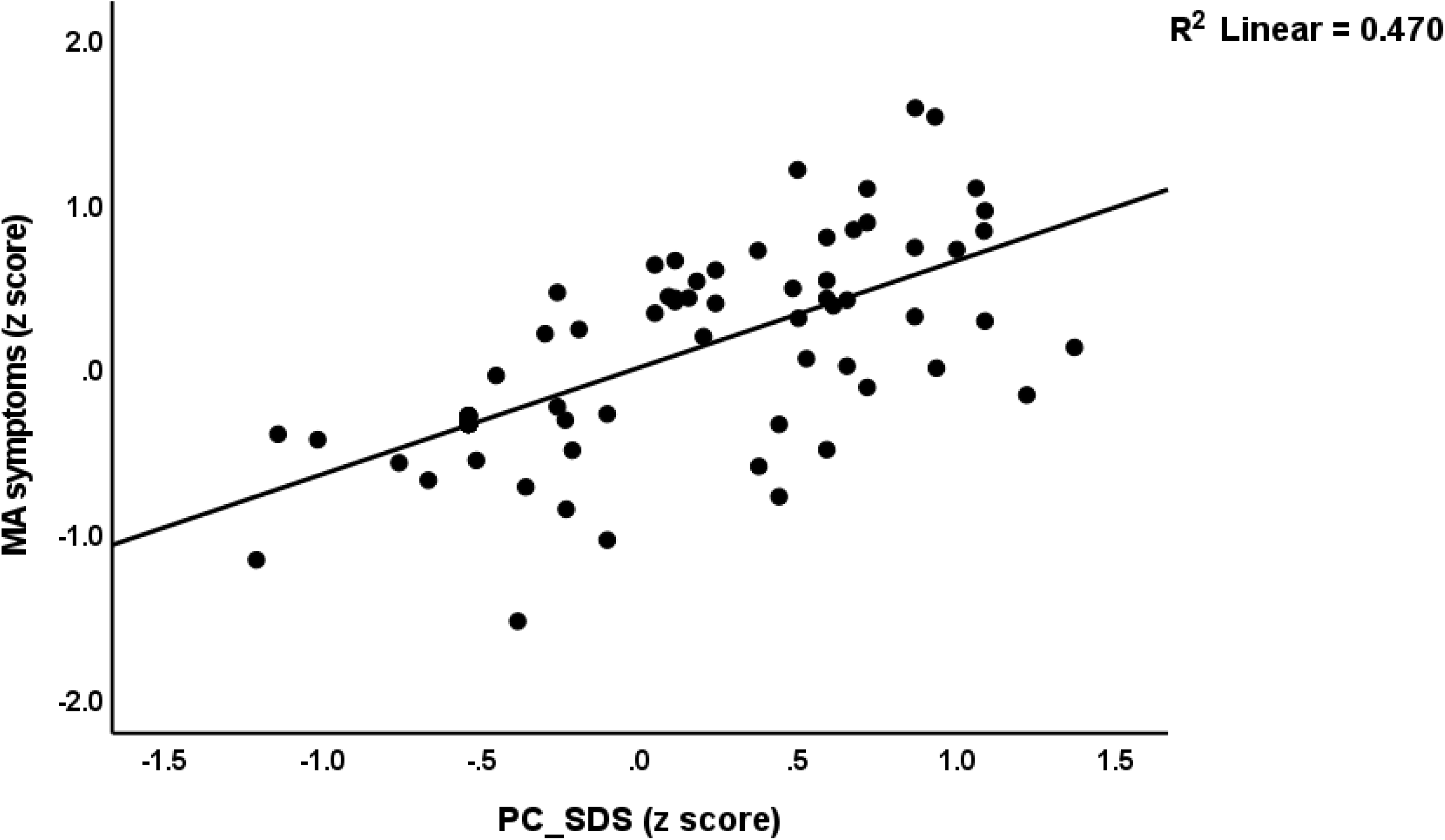
Partial regression plot of the overall severity of methamphetamine (MA)-induced psychotic symptoms (MA symptoms) on MA dependence (PC-SDS)

**Figure 5.**
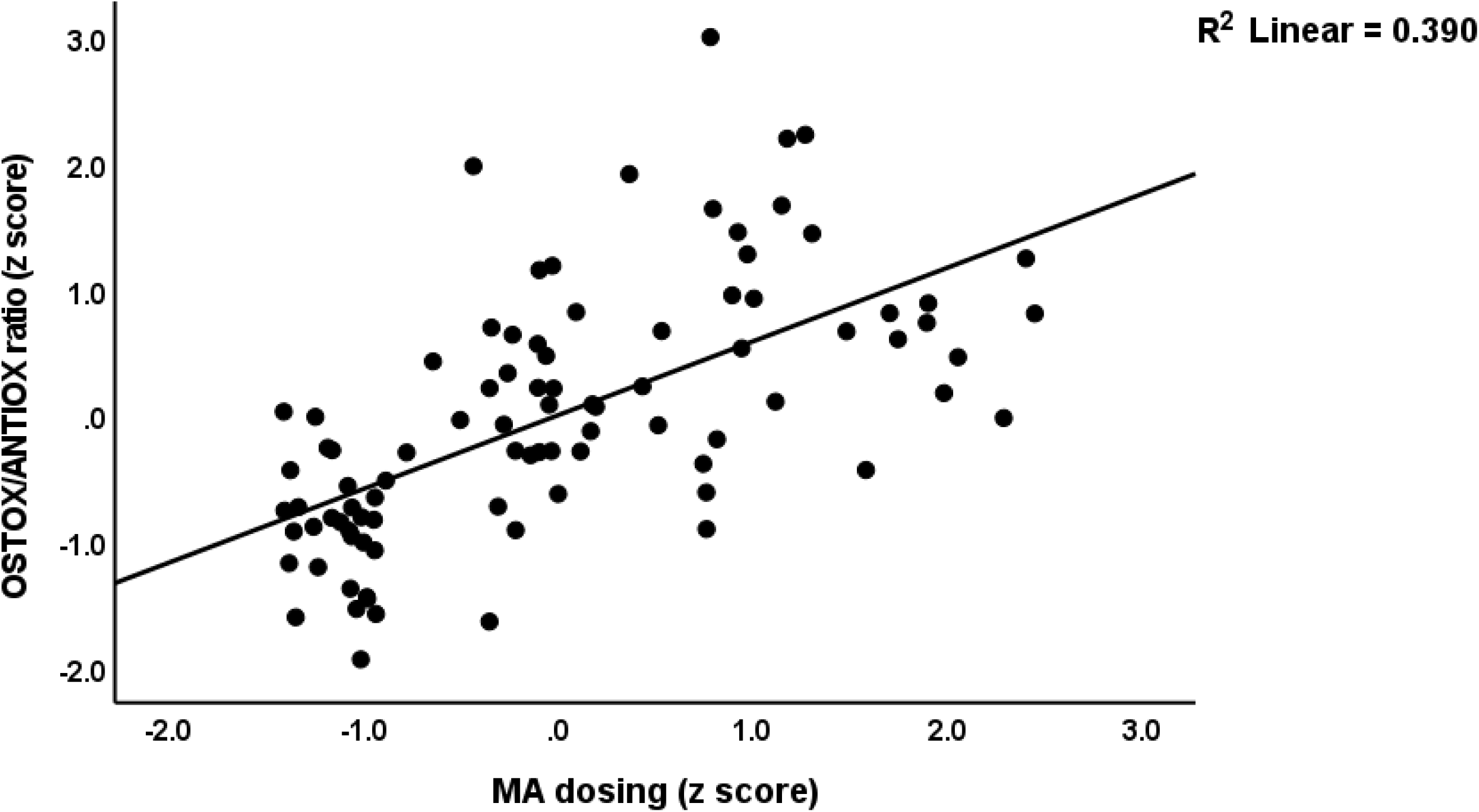
Partial regression plot of the oxidative toxicity / antioxidant defenses (OSTOX/ANTIOX) ratio on methamphetamine (MA) dosing

**Table 6.** Results of multiple regression analyses with methamphetamine (MA)-induced symptoms and the oxidative stress toxicity / antioxidant defenses (OSTOX/ANTIOX) ratio as dependent variables and MA dependence and MA intake features as explanatory variables

| Dependent Variables | Explanatory Variables | Parameter estimates + statistics |  |  | Model statistics and effect size |  |  |  |
| --- | --- | --- | --- | --- | --- | --- | --- | --- |
| | | $\beta$ | t | p | R<sup>2</sup> | F | df | p |
| #1 MA symptoms | **Model** |  |  |  | 0.818 | 194.96 | 2/87 | <0.001 |
|  | PC_SDS | 0.630 | 8.79 | <0.001 |  |  |  |  |
|  | MA dosing | 0.324 | 4.53 | <0.001 |  |  |  |  |
| #2. OSTOX/ANTIOX ratio | **Model** |  |  |  | 0.447 | 35.18 | 2/87 | <0.001 |
|  | PC_SDS | 0.449 | 3.60 | <0.001 |  |  |  |  |
|  | MA dosing | 0.258 | 2.07 | 0.041 |  |  |  |  |

PC\_SDS: a factor reflecting severity of MA dependence MA dosing: the dosing of MA prior to hospital admission.

### Results of PLS analysis

We used PLS analysis (**Figure 6**) to delineate whether the impact of MA use and dependence (entered as a latent vector extracted from MA use, MA dosing, route of administration, and PC_SDS) on the MA-related symptoms (entered as a latent vector extracted from five symptom domains (MAI symptoms, psychosis, hostility, excitation and FTD; mannerism did not load highly on this factor and was consequently deleted from the final model) is mediated by increased OSTOX and ANTIOX biomarkers. After feature reduction we found that HDL, oxDL, TAC and zinc were the significant predictors of the MIP symptoms. The quality of the current PLS model was adequate with SRMR = 0.042. The MA symptoms factor showed adequate convergence and construct reliability with AVE=0.853, rho A=0.967, composite reliability=0.970, and Cronbach alpha=0.957, while all loadings were > 0.887 at p < 0.001. The PC_MA symptoms factor also showed adequate convergence and construct reliability with AVE=0.756, rho A=0.914, composite reliability=0.924, and Cronbach alpha=0.886, while all loadings were > 0.665 at p<0.001. CTA confirmed that both latent vectors were not mis-specified as reflective models. PLSPredict showed that the construct indicators Q2 predict values were all > 0 indicating that the prediction error was lower than the naivest benchmark. Complete PLS analysis performed using 5,000 bootstraps revealed that 29.9% of the variance in MA symptoms could be explained by the regression on HDL, TAC, and zinc (all inversely) and oxLDL (positively). In addition, PC_MA predicted 10.3% of the variance in HDL, 13.2% in oxLDL, 20.2% in TAC and 15.2% in zinc. As such, PC_MA had significant specific indirect effects on MA symptoms mediated by oxLDL (t=2.24, p=0.013) and TAC (t=1.82, p=0.034) as well as significant total effects (t=5.15, p<0.001).

**Figure 6.**
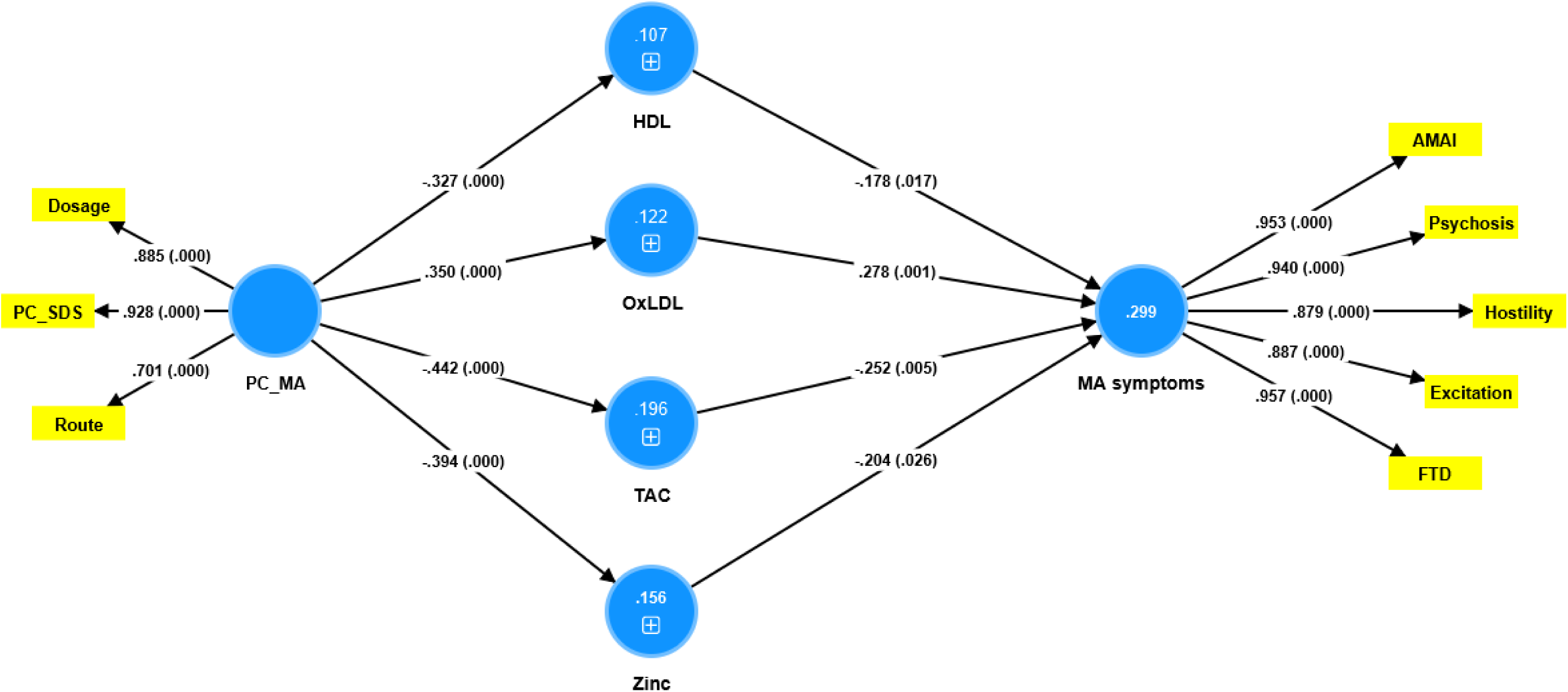
Results or Partial Least Squares (PLS) analysis which shows the impact of methamphetamine (MA) intake and dependence (PC_MA) on MA-induced symptoms which are partially mediated by increased levels of oxidized low-density lipoprotein (OxLDL), high-density lipoprotein (OxHDL) and lowered total antioxidant capacity (TAC) and zinc (Zn). MA symptoms are entered as a latent vector extracted from psychosis, hostility, excitation, mannerism, and MAI (acute MA-induced intoxication) symptoms. PC_MA is entered as a latent vector extracted from MA dosing, MA route of administration, and MA dependence (first principal component extracted from 4 SDS items). Shown are path coefficients (with exact p values between brackets), loadings (with p-values) of the latent vectors and the explained variances (white figures in blue circles).

## Discussion

### Clinical aspects of MA intoxication and MIP

The first major finding of the current study is that patients with acute intoxication could be divided into two relevant clusters: fifty percent of all patients could be assigned to a cluster with high scores on all PHEM symptoms and FTD as well as elevated OSTOX/ANTIOX values, and another 50% to a cluster with low symptoms and biomarker scores. In addition, the former group exhibit a greater severity of dependence, a longer duration of dependence, a higher MA dose prior to intoxication and hospitalization, and a greater proportion of MA injection rate. As such, a significant association was found between MA dependence/dosing/route, elevated OSTOX/ANTIOX levels, and psychotic symptoms.

Previous research revealed that 40% of MA users may exhibit positive psychotic symptoms and cognitive symptoms comparable to those found in schizophrenia, supporting that MIP may serve as a model for schizophrenia (Kalayasiri, Mutirangura et al. 2009, Glasner-Edwards and Mooney 2014, Hsieh, Stein et al. 2014, Kalayasiri, Verachai et al. 2014). Our findings also extend previous reports indicating that higher MA dependence, heavy MA use, increased frequency of use, and higher MA dosing (blood concentrations) are risk factors for MIP (review: (McKetin, Lubman et al. 2013, Arunogiri, Foulds et al. 2018).

MIP can be difficult to diagnose since it might be mistaken for a primary psychotic condition, such as schizophrenia or bipolar disorder, or a psychosis resulting from the use of another substance (Weich and Pienaar 2009, American Psychiatric 2013). In this respect, it should be highlighted that patients with premorbid schizophrenia or affective disorders were excluded from the present investigation and that no patients with other major illicit drug use disorders were included. Both MIP (this study) and schizophrenia (Maes, Plaimas et al. 2021, Maes, Vojdani et al. 2021, Al-Hakeim, Al-Musawi et al. 2022) are characterized by increased PHEM symptoms and FTD, although none of our patients suffered from clinically relevant hallucinations. However, many patients had high scores on delusions, conceptual disorganization, suspiciousness, excitement, and hostility. Previously it was described that MIP shares many symptomatic similarities with paranoid schizophrenia (e.g., based on DSM-IV-TR criteria) (McKetin, Lubman et al. 2013).

### MA dependence, OSTOX and ANTIOX

The second major finding of this study is that there are statistically significant differences in NOS biomarkers between patients with MA dependence and healthy controls. These findings extend those of earlier studies indicating that a substantial proportion of MA-dependent patients exhibit oxidative stress and psychosis as a result of their drug use (Zweben, Cohen et al. 2004, McKetin, McLaren et al. 2006, Bramness, Gundersen et al. 2012, Kalayasiri, Kraijak et al. 2019). In addition, we discovered that MA-dependent patients had significantly elevated MDA, oxHDL, oxLDL, OSTOX, and OSTOX/ANTIOX levels, and decreased catalase, TAC, HDL, zinc and ANTIOX levels as compared with controls. In addition, MA dependence coupled with MA dosing largely predicted increased OSTOX/ANTIOX values, with dependence being associated with OSTOX and MA dosing with decreased antioxidant defenses.

These findings extend previous findings that MA use increases MDA and other markers of lipid peroxidation in the blood and brain and decreases catalase, Gpx, GSH, SOD, and thiols groups (Mirecki, Fitzmaurice et al. 2004, Moszczynska, Fitzmaurice et al. 2004, Fitzmaurice, Tong et al. 2006, Govitrapong, Boontem et al. 2010, Suriyaprom, Tanateerabunjong et al. 2011, Huang, Lin et al. 2013, Hacimusalar, Karaaslan et al. 2019). MA also causes mitochondrial oxidative damage in human T lymphocytes (Potula, Hawkins et al. 2010) and in vivo and in vitro MA exposure increases ROS production in the CNS (Pubill, Chipana et al. 2005, Wu, Ping et al. 2007). Nonetheless, the present results demonstrate that in addition to MDA and Gpx, elevated oxLDL and oxHDL and decreased zinc and HDL are important alterations in MA dependence and abuse, and that it is essential to distinguish between MA dependence and MA dosing/route of administration.

Also, in rodent models, MA administration induces lipid peroxidation with elevated MDA and hydroxynonenal levels, protein oxidation with elevated protein carbonyl levels, increased NO and nitroprotein production, and decreased levels of antioxidant defenses including superoxide dismutase, catalase, and the glutathione system (McDonnell-Dowling and Kelly 2017). Prior animal studies demonstrated that chronic administration of MA causes nerve terminal degeneration in various brain regions, including the cortex, striatum, hippocampus, and olfactory bulb (Jeng, Ramkissoon et al. 2006, Deng, Ladenheim et al. 2007, Granado, Ares-Santos et al. 2011) and that these deleterious effects are mediated by ROS, including hydroxyl radicals, which cause oxidative damage not only to lipids but also to DNA and proteins (Jayanthi, Ladenheim et al. 1998, Jeng, Ramkissoon et al. 2006). When such damage cannot be repaired, it can accumulate and lead to cellular dysfunctions, neurotoxicity and neurodegeneration (Wells, McCallum et al. 2009, Moylan, Berk et al. 2014).

There is growing evidence that the neurotoxicity induced by MA is mediated by molecular pathways including immunological and oxidative processes, epigenetic changes, and changes in neurotransmitter turnover. MA use may cause the sustained release of catecholamines like dopamine (Kalayasiri, Kraijak et al. 2019, NIDA 2021), as well as neurodegeneration in the hippocampus and frontal, prefrontal, and temporal lobes; white matter hypertrophy and gliosis; glutamate neurotoxicity; and damage to neuronal dendrites (Thompson, Hayashi et al. 2004). MA-induced neurotoxicity is associated with increased production of dopaminergic quinones, increased production of ROS, inflammation and microglial activation, increased glutamatergic activity, activated apoptotic pathways, and activated oxidative pathways (Kita, Miyazaki et al. 2009, Yu, Zhu et al. 2015, Kalayasiri, Kraijak et al. 2019). The enhanced apoptotic pathways in MA-dependent patients, including BECN1, MAP1ALC3, CASP8, TP53, and BAX, may be explained by activated oxidative pathways (Zare, Ghanbari et al. 2021). Additionally,

MA may change the cholinergic anti-inflammatory system and the gut microbiota increasing leaky gut (Prakash, Tangalakis et al. 2017). Leaky gut may then further activate immune-inflammatory pathways and drive oxidative, immune-inflammatory, and related pathways. Additionally, modifications in LINE 1 partial methylation patterns caused by MA may result in changes in oxidative and immune-inflammatory pathways (Kalayasiri, Kraijak et al. 2019). MA exposure changes the production of not only immune-oxidative and apoptotic pathways but also neurotrophins, including brain-derived neurotrophic factor (BDNF) (Limanaqi, Gambardella et al. 2018, Eskandarian Boroujeni, Peirouvi et al. 2020). For example, long-term MA users had a substantial decrease in BDNF in the dorsolateral prefrontal cortex (Zare, Ghanbari et al. 2021). In animal studies, oxidative stress may decrease BDNF levels, whilst injection of vitamin E may reverse this impact (Wu, Ying et al. 2004). Importantly, tangled connections are reported between activated immunological and oxidative stress pathways and BDNF in major neuropsychiatric illnesses, with lower BDNF levels being related with diminished neurotrophic protection and subsequently greater neurotoxicity (Zare, Ghanbari et al. 2021, Mehterov, Minchev et al. 2022).

Nevertheless, our investigation revealed that MA dependence, acute intoxication, and MA dosage are accompanied by diminished antioxidant defenses. In this connection, Zare et al. (Zare, Ghanbari et al. 2021) showed a decreased GSH content in the dorsolateral prefrontal cortex of patients with persistent MA dependence and neurotoxic processes. Toborek et al. discovered that low levels of antioxidant defenses may contribute to the BBB damage induced by high levels of oxidative stress in MA-dependent individuals (Toborek, Seelbach et al. 2013).

It should be underscored that MA is accompanied by detrimental effects on cardiovascular health including atherosclerosis, coronary disease, and stroke (Kevil, Goeders et al. 2019, Schwarzbach, Lenk et al. 2020). Abuse of MA is known to enhance the production of atherosclerotic plaques and the risk of myocardial infarction (Kevil, Goeders et al. 2019). MA reduces some traditional risk factors, such as total cholesterol and BMI, but increases risk via effects on ROS and RNS production, proinflammatory genes and responses, and proatherogenic cytokines (including interleukin-1β and tumor necrosis factor-α), chemotaxis, adhesion molecules, endothelial activation, intimal cholesterol deposition, and increased catecholamine production (Kevil, Goeders et al. 2019, Schwarzbach, Lenk et al. 2020). Nonetheless, the present study demonstrates the existence of other and possibly even more significant proatherogenic risk factors, namely decreased HDL and elevated oxLDLand oxHDL levels. Oxidation of LDL is a significant early event in atherosclerotic processes via effects on lipoproteins, hypertension-related mechanisms, pro-inflammatory cytokines, and the production of IgG antibodies directed against oxLDL (Steinberg and Witztum 2010, Maes, Ruckoanich et al. 2011, Pirillo, Norata et al. 2013, Leiva, Wehinger et al. 2015). Increased production of oxHDL is a significant risk factor related with increased carotid intima-media thickness and cardiovascular mortality (Honda, Ueda et al. 2012). In addition, whereas HDL is a powerful antioxidant, anti-inflammatory, and anti-atherogenic molecule, oxHDL is dysfunctional, loses its antiatherogenic benefits, and may potentially become a proinflammatory compound that may even contribute to the advancement of atherosclerosis (Honda, Ueda et al. 2012, Matsunaga, Hara et al. 2014, Ito and Ito 2020).

Overall, there is compelling evidence that MA misuse and dependence produce damage due to oxidative stress and poorer antioxidant defenses, and that these processes play a crucial role in the enhanced neurotoxicity and atherosclerotic effects of MA.

### MA dependence, NOS biomarkers and MIP

The third main finding of this study is that changes in OSTOX and ANTIOX biomarkers predict MA-induced PHEM symptoms and FTD, and that lowered HDL, zinc, and TAC and increased oxHDL and oxLDL are the most important predictors of psychotic symptoms. In addition, we found that MA dependence/dosing is substantially linked with psychotic symptoms and that the effects of MA usage on MIP are partly mediated by increased OSTOX and decreased ANTIOX biomarkers. About 30% of the variance in PHEM symptoms and FTD may be explained by the cumulative effects of oxLDL, decreased HDL, TAC, and zinc. Consequently, the substantial effects of MA dependency and MA dosage on MIP may be partially explained by enhanced neurotoxicity due to impacts on several pathways, such as increased oxidative toxicity, as described in the preceding section. Importantly, neurotoxicity with increased lipid and protein oxidation and decreased antioxidant defenses and associated immune pathways plays a significant role in schizophrenia, particularly in the more severe phenotypes (Maes, Plaimas et al. 2021, Maes, Vojdani et al. 2021, Al-Hakeim, Al-Musawi et al. 2022). In both MIP and severe schizophrenia, the OSTOX/ANTIOX ratio is raised, and increased OSTOX and decreased ANTIOX partially predict PHEM symptoms and FTD. Intriguingly, both MIP and paranoid schizophrenia are characterized by abnormalities in partial LINE 1 methylation, with the latter being more prominent in paranoid schizophrenia (Kalayasiri, Kraijak et al. 2019). In addition, intertwined abnormalities in neuro-oxidative and neuro-immune pathways, and partial LINE 1 methylation may partially explain the pathogenesis of MIP and paranoid schizophrenia (Kalayasiri, Kraijak et al. 2019).

It is interesting to note that NO production was significantly decreased in the MA+PSO group and did not appear as a significant predictor of the symptoms in any of the multiple regression analyses. While increased NO production may have neurotoxic effects especially when oxygen radicals are increased, lowered levels of NO often indicate increased usage for formation of peroxynitrite, nitrosation and nitrosylation, which all have neurotoxic effects (Morris, Berk et al. 2017, Morris, Walder et al. 2018). Future research should examine these NO-associated pathways in MIP.

Overall, the findings suggest that elevated OSTOX and decreased ANTIOX during acute MA intoxication are associated with MIP and that increased oxidative toxicity and lowered antioxidant defenses are shared pathways between MIP and schizophrenia. Nevertheless, our findings that “only” 29.9% of the variance in MA psychotic symptoms was explained by oxidative and antioxidant biomarkers indicates that a larger part of the variance is determined by other pathways.

## Limitations

The present study would have been more interesting if we had examined the cytokine network, other oxidative stress biomarkers such as chlorinative stress, autoimmune responses to oxidative specific epitopes, and the tryptophan catabolite (TRYCAT) pathway, all of which play a role in schizophrenia (Maes, Vojdani et al. 2021, Almulla, Vasupanrajit et al. 2022). It could be argued that the sample size is rather small. Nevertheless, a priori power analysis showed that the minimum number of participants should be 82 to obtain a power of 0.8, whilst post hoc analysis shows that the obtained power was 0.98.

## Conclusions

PLS analysis revealed that HDL, TAC, and zinc (all inversely) and oxLDL (positively) explained 29.9% of the variance in MIP severity (a first factor extracted from psychosis, excitation, mannerism, and formal thought disorder scores). The severity of MA dependence and MA dosing and route of administration predict 10.3% of the variance in HDL, 13.2% in oxLDL, 20.2% in TAC, and 15.2% in zinc. MA dependency and intoxication are associated with elevated oxidative stress and diminished antioxidant defenses, both of which enhance the risk of MIP during acute intoxication.

## Data Availability

The dataset generated during and/or analyzed during the current study will be available from the corresponding author (M.M.) upon reasonable request and once the dataset has been fully exploited by the authors.

## Acknowledgments

The authors would like to thank the employees of the Psychiatry Unit at Al-Hussein Medical Center, Kerbala, Iraq for their hard work in compiling the data utilized in this study.

## Ethical approval and consent to participate

The study followed Iraqi and international privacy and ethics laws. Before participating in this study, all participants, first-degree relatives, or legal representatives. The study was approved by the ethics committee (IRB) of the College of Science, University of Kufa, Iraq (89/2022), Karbala Health Directorate-Training and Human Development Center (Document No.18378/ 2021), which follows the Declaration of Helsinki’s International Guideline for Human Research Protection.

## Author’s contributions

The recruitment of the participants and blood sampling were done by MFA. HKA and SRM assayed the biomarkers. AFA wrote the first draft of the manuscript. The statistical analysis was carried out by MM who also designed the methodology of the study. Final version of the manuscript is released following revising by all authors.

## References

Afzali, S., F. Fadaei, A. Oftadeh and A. Ranjbar (2022). “Salivary Biomarkers of Oxidative Stress in Methamphetamine Users: A Case-Control Study.” Novelty in Clinical Medicine 1(2): 95–100.

Al-Dujaili, A. H., R. F. Mousa, H. K. Al-Hakeim and M. Maes (2021). “High Mobility Group Protein 1 and Dickkopf-Related Protein 1 in Schizophrenia and Treatment-Resistant Schizophrenia: Associations With Interleukin-6, Symptom Domains, and Neurocognitive Impairments.” Schizophr Bull 47(2): 530–541.

Al-Hakeim, H. K., A. F. Al-Musawi, A. Al-Mulla, A. H. Al-Dujaili, M. Debnath and M. Maes (2022). “The interleukin-6/interleukin-23/T helper 17-axis as a driver of neuro-immune toxicity in the major neurocognitive psychosis or deficit schizophrenia: A precision nomothetic psychiatry analysis.” PLoS One 17(10): e0275839.

Al-Hakeim, H. K., A. F. Almulla and M. Maes (2020). “The Neuroimmune and Neurotoxic Fingerprint of Major Neurocognitive Psychosis or Deficit Schizophrenia: a Supervised Machine Learning Study.” Neurotoxicity Research 37(3): 753–771.

Almulla, A. F., H. K. Al-Hakeim and M. Maes (2021). “Schizophrenia phenomenology revisited: positive and negative symptoms are strongly related reflective manifestations of an underlying single trait indicating overall severity of schizophrenia.” CNS Spectr 26(4): 368–377.

Almulla, A. F., A. Vasupanrajit, C. Tunvirachaisakul, H. K. Al-Hakeim, M. Solmi, R. Verkerk and M. Maes (2022). “The tryptophan catabolite or kynurenine pathway in schizophrenia: meta-analysis reveals dissociations between central, serum, and plasma compartments.” Molecular Psychiatry.

American Psychiatric, A. (2013). Diagnostic and Statistical Manual of Mental Disorders.

American Psychiatric Association, A. (2013). Diagnostic and statistical manual of mental disorders.

Arunogiri, S., J. A. Foulds, R. McKetin and D. I. Lubman (2018). “A systematic review of risk factors for methamphetamine-associated psychosis.” Australian & New Zealand Journal of Psychiatry 52(6): 514–529.

Batki, S. L. and D. S. Harris (2004). “Quantitative drug levels in stimulant psychosis: relationship to symptom severity, catecholamines and hyperkinesia.” Am J Addict 13(5): 461–470.

Beck, A. K., B. Larance, V. Manning, L. Hides, A. L. Baker, F. P. Deane, A. Shakeshaft, D. Raftery and P. J. Kelly (2022). “Online SMART Recovery mutual support groups: Characteristics and experience of adults seeking treatment for methamphetamine compared to those seeking treatment for other addictive behaviours.” Drug Alcohol Rev.

Benjamini, Y. and Y. Hochberg (1995). “Controlling the False Discovery Rate: A Practical and Powerful Approach to Multiple Testing.” Journal of the Royal Statistical Society: Series B (Methodological) 57(1): 289–300.

Boll, K. M., C. Noto, K. L. Bonifácio, C. C. Bortolasci, A. Gadelha, R. A. Bressan, D. S. Barbosa, M. Maes and E. G. Moreira (2017). “Oxidative and nitrosative stress biomarkers in chronic schizophrenia.” Psychiatry Res 253: 43–48.

Bramness, J. G., Ø. H. Gundersen, J. Guterstam, E. B. Rognli, M. Konstenius, E.-M. Løberg, S. Medhus, L. Tanum and J. Franck (2012). “Amphetamine-induced psychosis - a separate diagnostic entity or primary psychosis triggered in the vulnerable?” BMC Psychiatry 12(1): 221.

Cuenod, M., P. Steullet, J.-H. Cabungcal, D. Dwir, I. Khadimallah, P. Klauser, P. Conus and K. Q. Do (2022). “Caught in vicious circles: a perspective on dynamic feed-forward loops driving oxidative stress in schizophrenia.” Molecular Psychiatry 27(4): 1886–1897.

Deng, X., B. Ladenheim, S. Jayanthi and J. L. Cadet (2007). “Methamphetamine Administration Causes Death of Dopaminergic Neurons in the Mouse Olfactory Bulb.” Biological Psychiatry 61(11): 1235–1243.

Ermakov, E. A., E. M. Dmitrieva, D. A. Parshukova, D. V. Kazantseva, A. R. Vasilieva and L. P. Smirnova (2021). “Oxidative Stress-Related Mechanisms in Schizophrenia Pathogenesis and New Treatment Perspectives.” Oxid Med Cell Longev 2021: 8881770.

Eskandarian Boroujeni, M., T. Peirouvi, F. Shaerzadeh, A. Ahmadiani, M. A. Abdollahifar and A. Aliaghaei (2020). “Differential gene expression and stereological analyses of the cerebellum following methamphetamine exposure.” Addict Biol 25(1): e12707.

Fitzmaurice, P. S., J. Tong, M. Yazdanpanah, P. P. Liu, K. S. Kalasinsky and S. J. Kish (2006). “Levels of 4-hydroxynonenal and malondialdehyde are increased in brain of human chronic users of methamphetamine.” J Pharmacol Exp Ther 319(2): 703–709.

Gan, H., Z. Song, P. Xu, H. Su, Y. Pan, M. Zhao and D. Liu (2018). “A comparison study of working memory deficits between patients with methamphetamine-associated psychosis and patients with schizophrenia.” Shanghai Archives of Psychiatry 30(3): 168.

Glasner-Edwards, S. and L. J. Mooney (2014). “Methamphetamine Psychosis: Epidemiology and Management.” CNS Drugs 28(12): 1115–1126.

Glasner-Edwards, S., L. J. Mooney, P. Marinelli-Casey, M. Hillhouse, A. Ang and R. Rawson (2008). “Clinical course and outcomes of methamphetamine-dependent adults with psychosis.” J Subst Abuse Treat 35(4): 445–450.

Glasner-Edwards, S., L. J. Mooney, P. Marinelli-Casey, M. Hillhouse, A. Ang and R. A. Rawson (2010). “Psychopathology in methamphetamine-dependent adults 3 years after treatment.” Drug Alcohol Rev 29(1): 12–20.

Gossop, M., S. Darke, P. Griffiths, J. Hando, B. Powis, W. Hall and J. Strang (1995). “The Severity of Dependence Scale (SDS): psychometric properties of the SDS in English and Australian samples of heroin, cocaine and amphetamine users.” Addiction 90(5): 607–614.

Govitrapong, P., P. Boontem, P. Kooncumchoo, S. Pinweha, J. Namyen, Y. Sanvarinda and S. Vatanatunyakum (2010). “Increased blood oxidative stress in amphetamine users.” Addict Biol 15(1): 100–102.

Granado, N., S. Ares-Santos, I. Oliva, E. O’Shea, E. D. Martin, M. I. Colado and R. Moratalla (2011). “Dopamine D2-receptor knockout mice are protected against dopaminergic neurotoxicity induced by methamphetamine or MDMA.” Neurobiol Dis 42(3): 391–403.

Guidara, W., M. Messedi, M. Naifar, M. Maalej, S. Grayaa, S. Omri, J. B. Thabet, M. Maalej, N. Charfi and F. Ayadi (2020). “Predictive value of oxidative stress biomarkers in drug-free patients with schizophrenia and schizo-affective disorder.” Psychiatry research 293: 113467.

Hacimusalar, Y., O. Karaaslan, C. Bal, D. Kocer, G. Gok and B. Yildiz (2019). “Methamphetamine’s effects on oxidative stress markers may continue after detoxification: a case–control study.” Psychiatry and Clinical Psychopharmacology 29(3): 361–367.

Harro, J. (2015). “Neuropsychiatric Adverse Effects of Amphetamine and Methamphetamine.” Int Rev Neurobiol 120: 179–204.

Hogarth, S., E. Manning and M. van den Buuse (2021). Chronic Methamphetamine and Psychosis Pathways. <u>Handbook of Substance Misuse and Addictions: From Biology to Public Health</u>. V. B. Patel and V. R. Preedy. Cham, Springer International Publishing: 1-26.

Honda, H., M. Ueda, S. Kojima, S. Mashiba, T. Michihata, K. Takahashi, K. Shishido and T. Akizawa (2012). “Oxidized high-density lipoprotein as a risk factor for cardiovascular events in prevalent hemodialysis patients.” Atherosclerosis 220(2): 493–501.

Hsieh, J. H., D. J. Stein and F. M. Howells (2014). “The neurobiology of methamphetamine induced psychosis.” Frontiers in Human Neuroscience 8.

Huang, M.-C., S.-K. Lin, C.-H. Chen, C.-H. Pan, C.-H. Lee and H.-C. Liu (2013). “Oxidative stress status in recently abstinent methamphetamine abusers.” Psychiatry and Clinical Neurosciences 67(2): 92–100.

Ikeda, M., Y. Okahisa, B. Aleksic, M. Won, N. Kondo, N. Naruse, K. Aoyama-Uehara, I. Sora, M. Iyo and R. Hashimoto (2013). “Evidence for shared genetic risk between methamphetamine-induced psychosis and schizophrenia.” Neuropsychopharmacology 38(10): 1864–1870.

Ito, F. and T. Ito (2020) “High-Density Lipoprotein (HDL) Triglyceride and Oxidized HDL: New Lipid Biomarkers of Lipoprotein-Related Atherosclerotic Cardiovascular Disease.” Antioxidants 9 DOI: 10.3390/antiox9050362.

Jayanthi, S., B. Ladenheim and J. L. Cadet (1998). “Methamphetamine-induced changes in antioxidant enzymes and lipid peroxidation in copper/zinc-superoxide dismutase transgenic mice.” Ann N Y Acad Sci 844: 92–102.

Jeng, W., A. Ramkissoon, T. Parman and P. G. Wells (2006). “Prostaglandin H synthase-catalyzed bioactivation of amphetamines to free radical intermediates that cause CNS regional DNA oxidation and nerve terminal degeneration.” Faseb j 20(6): 638–650.

Jeng, W., A. Ramkissoon, T. Parman and P. G. Wells (2006). “Prostaglandin H synthase-catalyzed bioactivation of amphetamines to free radical intermediates that cause CNS regional DNA oxidation and nerve terminal degeneration1.” The FASEB Journal 20(6): 638–650.

Jones, C. M., D. Houry, B. Han, G. Baldwin, A. Vivolo-Kantor and W. M. Compton (2022). “Methamphetamine use in the United States: epidemiological update and implications for prevention, treatment, and harm reduction.” Annals of the New York Academy of Sciences 1508(1): 3–22.

Kalayasiri, R., K. Kraijak, M. Maes and A. Mutirangura (2019). “Methamphetamine (MA) Use Induces Specific Changes in LINE-1 Partial Methylation Patterns, Which Are Associated with MA-Induced Paranoia: a Multivariate and Neuronal Network Study.” Mol Neurobiol 56(6): 4258–4272.

Kalayasiri, R., K. Kraijak, A. Mutirangura and M. Maes (2019). “Paranoid schizophrenia and methamphetamine-induced paranoia are both characterized by a similar LINE-1 partial methylation profile, which is more pronounced in paranoid schizophrenia.” Schizophrenia Research 208: 221–227.

Kalayasiri, R., A. Mutirangura, V. Verachai, J. Gelernter and R. T. Malison (2009). “Risk factors for methamphetamine-induced paranoia and latency of symptom onset in a Thai drug treatment cohort.” Asian Biomedicine 3(6): 635–643.

Kalayasiri, R., V. Verachai, J. Gelernter, A. Mutirangura and R. T. Malison (2014). “Clinical features of methamphetamine-induced paranoia and preliminary genetic association with DBH-1021C→T in a Thai treatment cohort.” Addiction 109(6): 965–976.

Kay, S. R., A. Fiszbein and L. A. Opler (1987). “The positive and negative syndrome scale (PANSS) for schizophrenia.” Schizophrenia bulletin 13(2): 261–276.

Kevil, C. G., N. E. Goeders, M. D. Woolard, M. S. Bhuiyan, P. Dominic, G. K. Kolluru, C. L. Arnold, J. G. Traylor and A. W. Orr (2019). “Methamphetamine Use and Cardiovascular Disease.” Arteriosclerosis, Thrombosis, and Vascular Biology 39(9): 1739–1746.

Kita, T., I. Miyazaki, M. Asanuma, M. Takeshima and G. C. Wagner (2009). Chapter 3 - Dopamine-Induced Behavioral Changes and Oxidative Stress in Methamphetamine-Induced Neurotoxicity. <u>International Review of Neurobiology</u>, Academic Press. 88: 43-64.

Lecomte, T., A. Dumais, J. R. Dugre and S. Potvin (2018). “The prevalence of substance-induced psychotic disorder in methamphetamine misusers: a meta-analysis.” Psychiatry research 268: 189–192.

Leiva, E., S. Wehinger, L. Guzmán and R. Orrego (2015). “Role of oxidized LDL in atherosclerosis.” Hypercholesterolemia: 55–78.

Limanaqi, F., S. Gambardella, F. Biagioni, C. L. Busceti and F. Fornai (2018). “Epigenetic effects induced by methamphetamine and methamphetamine-dependent oxidative stress.” Oxidative medicine and cellular longevity 2018.

Limanaqi, F., S. Gambardella, F. Biagioni, C. L. Busceti and F. Fornai (2018). “Epigenetic Effects Induced by Methamphetamine and Methamphetamine-Dependent Oxidative Stress.” Oxid Med Cell Longev 2018: 4982453.

Maes, M. (2022). “Precision Nomothetic Medicine in Depression Research: A New Depression Model, and New Endophenotype Classes and Pathway Phenotypes, and A Digital Self.” J Pers Med 12(3).

Maes, M., K. Plaimas, A. Suratanee, C. Noto and B. Kanchanatawan (2021). “First Episode Psychosis and Schizophrenia Are Systemic Neuro-Immune Disorders Triggered by a Biotic Stimulus in Individuals with Reduced Immune Regulation and Neuroprotection.” Cells 10(11).

Maes, M., P. Ruckoanich, Y. S. Chang, N. Mahanonda and M. Berk (2011). “Multiple aberrations in shared inflammatory and oxidative & nitrosative stress (IO&NS) pathways explain the co-association of depression and cardiovascular disorder (CVD), and the increased risk for CVD and due mortality in depressed patients.” Prog Neuropsychopharmacol Biol Psychiatry 35(3): 769–783.

Maes, M., S. Sirivichayakul, A. K. Matsumoto, A. Maes, A. P. Michelin, L. de Oliveira Semeão, J. V. de Lima Pedrão, E. G. Moreira, D. S. Barbosa, M. Geffard, A. F. Carvalho and B. Kanchanatawan (2020). “Increased Levels of Plasma Tumor Necrosis Factor-α Mediate Schizophrenia Symptom Dimensions and Neurocognitive Impairments and Are Inversely Associated with Natural IgM Directed to Malondialdehyde and Paraoxonase 1 Activity.” Mol Neurobiol 57(5): 2333–2345.

Maes, M., S. Sirivichayakul, A. K. Matsumoto, A. P. Michelin, L. de Oliveira Semeão, J. V. de Lima Pedrão, E. G. Moreira, D. S. Barbosa, A. F. Carvalho, M. Solmi and B. Kanchanatawan (2020). “Lowered Antioxidant Defenses and Increased Oxidative Toxicity Are Hallmarks of Deficit Schizophrenia: a Nomothetic Network Psychiatry Approach.” Molecular Neurobiology 57(11): 4578–4597.

Maes, M., A. Vojdani, S. Sirivichayakul, D. S. Barbosa and B. Kanchanatawan (2021). “Inflammatory and Oxidative Pathways Are New Drug Targets in Multiple Episode Schizophrenia and Leaky Gut, Klebsiella pneumoniae, and C1q Immune Complexes Are Additional Drug Targets in First Episode Schizophrenia.” Mol Neurobiol 58(7): 3319–3334.

Matsunaga, T., A. Hara and T. Komoda (2014). Chapter 10 - Oxidized High-Density Lipoprotein: Friend or Foe. <u>The HDL Handbook (Second Edition)</u>. T. Komoda. Boston, Academic Press: 247–272.

May, A. C., R. L. Aupperle and J. L. Stewart (2020). “Dark Times: The Role of Negative Reinforcement in Methamphetamine Addiction.” Front Psychiatry 11: 114.

McDonnell-Dowling, K. and J. P. Kelly (2017). “The Role of Oxidative Stress in Methamphetamine-induced Toxicity and Sources of Variation in the Design of Animal Studies.” Curr Neuropharmacol 15(2): 300–314.

McKetin, R. (2018). “Methamphetamine psychosis: insights from the past.” Addiction 113(8): 1522–1527.

McKetin, R., D. I. Lubman, A. L. Baker, S. Dawe and R. L. Ali (2013). “Dose-related psychotic symptoms in chronic methamphetamine users: evidence from a prospective longitudinal study.” JAMA Psychiatry 70(3): 319–324.

McKetin, R., D. I. Lubman, J. M. Najman, S. Dawe, P. Butterworth and A. L. Baker (2014). “Does methamphetamine use increase violent behaviour? Evidence from a prospective longitudinal study.” Addiction 109(5): 798–806.

McKetin, R., J. McLaren, D. I. Lubman and L. Hides (2006). “The prevalence of psychotic symptoms among methamphetamine users.” Addiction 101(10): 1473–1478.

Mehterov, N., D. Minchev, M. Gevezova, V. Sarafian and M. Maes (2022). “Interactions Among Brain-Derived Neurotrophic Factor and Neuroimmune Pathways Are Key Components of the Major Psychiatric Disorders.” Mol Neurobiol 59(8): 4926–4952.

Meredith, C. W., C. Jaffe, K. Ang-Lee and A. J. Saxon (2005). “Implications of chronic methamphetamine use: a literature review.” Harv Rev Psychiatry 13(3): 141–154.

Mirecki, A., P. Fitzmaurice, L. Ang, K. S. Kalasinsky, F. J. Peretti, S. S. Aiken, D. J. Wickham, A. Sherwin, J. N. Nobrega, H. J. Forman and S. J. Kish (2004). “Brain antioxidant systems in human methamphetamine users.” J Neurochem 89(6): 1396–1408.

Morris, G., M. Berk, H. Klein, K. Walder, P. Galecki and M. Maes (2017). “Nitrosative Stress, Hypernitrosylation, and Autoimmune Responses to Nitrosylated Proteins: New Pathways in Neuroprogressive Disorders Including Depression and Chronic Fatigue Syndrome.” Mol Neurobiol 54(6): 4271–4291.

Morris, G., K. Walder, A. F. Carvalho, S. J. Tye, K. Lucas, M. Berk and M. Maes (2018). “The role of hypernitrosylation in the pathogenesis and pathophysiology of neuroprogressive diseases.” Neurosci Biobehav Rev 84: 453–469.

Moszczynska, A., P. Fitzmaurice, L. Ang, K. S. Kalasinsky, F. J. Peretti, S. S. Aiken, D. J. Wickham, A. Sherwin, J. N. Nobrega, H. J. Forman and S. J. Kish (2004). “Brain antioxidant systems in human methamphetamine users.” Journal of Neurochemistry 89(6): 1396–1408.

Moylan, S., M. Berk, O. M. Dean, Y. Samuni, L. J. Williams, A. O’Neil, A. C. Hayley, J. A. Pasco, G. Anderson, F. N. Jacka and M. Maes (2014). “Oxidative & nitrosative stress in depression: why so much stress?” Neurosci Biobehav Rev 45: 46–62.

NIDA. (2021). “ Overview “, Accessed October 3, 2022.

NIDA (2022). What are the long-term effects of methamphetamine misuse? Retrieved from https://nida.nih.gov/publications/research-reports/methamphetamine/what-are-long-term-effects-methamphetamine-misuse on 2022, September 19.

Noto, C., M. Maes, V. K. Ota, A. L. Teixeira, R. A. Bressan, A. Gadelha and E. Brietzke (2015). “High predictive value of immune-inflammatory biomarkers for schizophrenia diagnosis and association with treatment resistance.” The World Journal of Biological Psychiatry 16(6): 422–429.

Overall, J. E. and D. R. Gorham (1962). “The brief psychiatric rating scale.” Psychological reports 10(3): 799–812.

Pirillo, A., G. D. Norata and A. L. Catapano (2013). “LOX-1, OxLDL, and atherosclerosis.” Mediators Inflamm 2013: 152786.

Potula, R., B. J. Hawkins, J. M. Cenna, S. Fan, H. Dykstra, S. H. Ramirez, B. Morsey, M. R. Brodie and Y. Persidsky (2010). “Methamphetamine causes mitrochondrial oxidative damage in human T lymphocytes leading to functional impairment.” J Immunol 185(5): 2867–2876.

Prakash, M. D., K. Tangalakis, J. Antonipillai, L. Stojanovska, K. Nurgali and V. Apostolopoulos (2017). “Methamphetamine: Effects on the brain, gut and immune system.” Pharmacological Research 120: 60–67.

Pubill, D., C. Chipana, A. Camins, M. Pallàs, J. Camarasa and E. Escubedo (2005). “Free radical production induced by methamphetamine in rat striatal synaptosomes.” Toxicol Appl Pharmacol 204(1): 57–68.

Roomruangwong, C., C. Noto, B. Kanchanatawan, G. Anderson, M. Kubera, A. F. Carvalho and M. Maes (2020). “The Role of Aberrations in the Immune-Inflammatory Response System (IRS) and the Compensatory Immune-Regulatory Reflex System (CIRS) in Different Phenotypes of Schizophrenia: the IRS-CIRS Theory of Schizophrenia.” Molecular Neurobiology 57(2): 778–797.

Russell, K., D. M. Dryden, Y. Liang, C. Friesen, K. O’Gorman, T. Durec, T. C. Wild and T. P. Klassen (2008). “Risk factors for methamphetamine use in youth: a systematic review.” BMC Pediatrics 8(1): 48.

Salo, R., C. Fassbender, A.-M. Iosif, S. Ursu, M. H. Leamon and C. Carter (2013). “Predictors of methamphetamine psychosis: History of ADHD-relevant childhood behaviors and drug exposure.” Psychiatry research 210(2): 529–535.

Schwarzbach, V., K. Lenk and U. Laufs (2020). “Methamphetamine-related cardiovascular diseases.” ESC Heart Failure 7(2): 407–414.

Shoptaw, S., M. J. Li, M. Javanbakht, A. Ragsdale, D. Goodman-Meza and P. M. Gorbach (2022). “Frequency of reported methamphetamine use linked to prevalence of clinical conditions, sexual risk behaviors, and social adversity in diverse men who have sex with men in Los Angeles.” Drug and Alcohol Dependence 232: 109320.

Solberg, D. K., H. Refsum, O. A. Andreassen and H. Bentsen (2019). “A five-year follow-up study of antioxidants, oxidative stress and polyunsaturated fatty acids in schizophrenia.” Acta neuropsychiatrica 31(4): 202–212.

Steinberg, D. and J. L. Witztum (2010). “Oxidized low-density lipoprotein and atherosclerosis.” Arterioscler Thromb Vasc Biol 30(12): 2311–2316.

Strickland, J. C., W. W. Stoops, K. E. Dunn, K. E. Smith and J. R. Havens (2021). “The continued rise of methamphetamine use among people who use heroin in the United States.” Drug and Alcohol Dependence 225: 108750.

Suriyaprom, K., R. Tanateerabunjong, A. Tungtrongchitr and R. Tungtrongchitr (2011). “Alterations in malondialdehyde levels and laboratory parameters among methamphetamine abusers.” J Med Assoc Thai 94(12): 1533–1539.

Thompson, P. M., K. M. Hayashi, S. L. Simon, J. A. Geaga, M. S. Hong, Y. Sui, J. Y. Lee, A. W. Toga, W. Ling and E. D. London (2004). “Structural Abnormalities in the Brains of Human Subjects Who Use Methamphetamine.” The Journal of Neuroscience 24(26): 6028.

Toborek, M., M. J. Seelbach, C. S. Rashid, I. E. András, L. Chen, M. Park and K. A. Esser (2013). “Voluntary exercise protects against methamphetamine-induced oxidative stress in brain microvasculature and disruption of the blood–brain barrier.” Molecular Neurodegeneration 8(1): 22.

Wearne, T. A. and J. L. Cornish (2018). “A Comparison of Methamphetamine-Induced Psychosis and Schizophrenia: A Review of Positive, Negative, and Cognitive Symptomatology.” Frontiers in Psychiatry 9.

Weich, L. and W. Pienaar (2009). “Occurrence of comorbid substance use disorders among acute psychiatric inpatients at Stikland Hospital in the Western Cape, South Africa.” Afr J Psychiatry (Johannesbg) 12(3): 213–217.

Wells, P. G., G. P. McCallum, C. S. Chen, J. T. Henderson, C. J. J. Lee, J. Perstin, T. J. Preston, M. J. Wiley and A. W. Wong (2009). “Oxidative Stress in Developmental Origins of Disease: Teratogenesis, Neurodevelopmental Deficits, and Cancer.” Toxicological Sciences 108(1): 4–18.

Wu, A., Z. Ying and F. Gomez-Pinilla (2004). “The interplay between oxidative stress and brain-derived neurotrophic factor modulates the outcome of a saturated fat diet on synaptic plasticity and cognition.” Eur J Neurosci 19(7): 1699–1707.

Wu, C. W., Y. H. Ping, J. C. Yen, C. Y. Chang, S. F. Wang, C. L. Yeh, C. W. Chi and H. C. Lee (2007). “Enhanced oxidative stress and aberrant mitochondrial biogenesis in human neuroblastoma SH-SY5Y cells during methamphetamine induced apoptosis.” Toxicol Appl Pharmacol 220(3): 243–251.

Yu, S., L. Zhu, Q. Shen, X. Bai and X. Di (2015). “Recent advances in methamphetamine neurotoxicity mechanisms and its molecular pathophysiology.” Behav Neurol 2015: 103969.

Zare, A., A. Ghanbari, M. J. Hoseinpour, M. Eskandarian Boroujeni, A. Alimohammadi, M. A. Abdollahifar, A. Aliaghaei, V. Mansouri and H. Z. Arani (2021). “Methamphetamine-Triggered Neurotoxicity in Human Dorsolateral Prefrontal Cortex.” Galen Med J 10: e2016.

Zweben, J. E., J. B. Cohen, D. Christian, G. P. Galloway, M. Salinardi, D. Parent and M. Iguchi (2004). “Psychiatric symptoms in methamphetamine users.” Am J Addict 13(2): 181–190.

Zweben, J. E., J. B. Cohen, D. Christian, G. P. Galloway, M. Salinardi, D. Parent, M. Iguchi and P. Methamphetamine Treatment (2004). “Psychiatric Symptoms in Methamphetamine Users.” The American Journal on Addictions 13(2): 181–190.

